# Single cell variant to enhancer to gene map for coronary artery disease

**DOI:** 10.1101/2024.11.13.24317257

**Authors:** Junedh M. Amrute, Paul C. Lee, Ittai Eres, Chang Jie Mick Lee, Andrea Bredemeyer, Maya U. Sheth, Tracy Yamawaki, Rijan Gurung, Chukwuemeka Anene-Nzelu, Wei-Lin Qiu, Soumya Kundu, Daniel Y. Li, Markus Ramste, Daniel Lu, Anthony Tan, Chul-Joo Kang, Ryan E. Wagoner, Arturo Alisio, Paul Cheng, Quanyi Zhao, Clint L. Miller, Ira M. Hall, Rajat M. Gupta, Yi-Hsiang Hsu, Saptarsi M. Haldar, Kory J. Lavine, Simon Jackson, Robin Andersson, Jesse M. Engreitz, Roger S-Y Foo, Chi-Ming Li, Brandon Ason, Thomas Quertermous, Nathan O. Stitziel

## Abstract

Although genome wide association studies (GWAS) in large populations have identified hundreds of variants associated with common diseases such as coronary artery disease (CAD), most disease-associated variants lie within non-coding regions of the genome, rendering it difficult to determine the downstream causal gene and cell type. Here, we performed paired single nucleus gene expression and chromatin accessibility profiling from 44 human coronary arteries. To link disease variants to molecular traits, we developed a meta-map of 88 samples and discovered 11,182 single-cell chromatin accessibility quantitative trait loci (caQTLs). Heritability enrichment analysis and disease variant mapping demonstrated that smooth muscle cells (SMCs) harbor the greatest genetic risk for CAD. To capture the continuum of SMC cell states in disease, we used dynamic single cell caQTL modeling for the first time in tissue to uncover QTLs whose effects are modified by cell state and expand our insight into genetic regulation of heterogenous cell populations. Notably, we identified a variant in the *COL4A1*/*COL4A2* CAD GWAS locus which becomes a caQTL as SMCs de-differentiate by changing a transcription factor binding site for EGR1/2. To unbiasedly prioritize functional candidate genes, we built a genome-wide single cell variant to enhancer to gene (scV2E2G) map for human CAD to link disease variants to causal genes in cell types. Using this approach, we found several hundred genes predicted to be linked to disease variants in different cell types. Next, we performed genome-wide Hi-C in 16 human coronary arteries to build tissue specific maps of chromatin conformation and link disease variants to integrated chromatin hubs and distal target genes. Using this approach, we show that rs4887091 within the *ADAMTS7* CAD GWAS locus modulates function of a super chromatin interactome through a change in a CTCF binding site. Finally, we used CRISPR interference to validate a distal gene, *AMOTL2*, liked to a CAD GWAS locus. Collectively we provide a disease-agnostic framework to translate human genetic findings to identify pathologic cell states and genes driving disease, producing a comprehensive scV2E2G map with genetic and tissue level convergence for future mechanistic and therapeutic studies.

## Introduction

Common diseases are driven by a multitude of factors and are among the most difficult to treat given the lack of a unified disease mechanism^1,2^. Coronary artery disease (CAD) is the most prevalent common heart disease and a leading cause of death worldwide^3^. Numerous genome-wide association studies (GWAS) have uncovered causal disease variants driving CAD risk and paved the way for new therapeutic target discovery^4–9^. Notably, much of the translation from GWAS to therapeutic targets in the CAD space has been restricted to lipid-lowering drugs^10,11^. While classically considered a lipid driven disease, large-scale clinical outcome studies have shown a persistent risk of CAD independent of lipid levels and patients continue to suffer from atherosclerotic disease despite optimal lipid-lowering therapy^12–14^. Additionally, most of the GWAS loci in CAD are thought to regulate biological pathways outside lipid biology^15,16^. Leveraging human genetics to uncover new lipid-independent biological mechanisms and drug targets in CAD presents a tremendous unmet need and would be a major milestone in translating population-scale GWAS findings to therapies for patients^17–19^.

While population scale GWAS studies offer the greatest opportunity for target discovery, most of the variants identified lie within non-coding regions of the genome, making it difficult to dissect the causal cells and molecular pathways^20,21^. The advent of single cell sequencing technologies now allows for the profiling of human tissues in health and disease at unprecedented granularity^22–25^. Prior studies have utilized traditional single cell RNA-sequencing (scRNA-seq) for detailed characterization of cell states in human tissue that drive disease^26–33^; however, these studies do not profile the non-coding genome making it challenging to link GWAS variants to observed molecular changes. More recently, Turner *et al*^34^ utilized single-nucleus assay for transposase-accessible chromatin with sequencing (snATAC-seq) in human coronary arteries to link CAD GWAS variants to regulatory elements. Although this represented a major advance in the field, the lack of complementary transcriptomic profiling rendered it challenging to nominate the causal genes in these specific cell types. Furthermore, isolated snATAC-seq precludes the discovery of distal gene regulation as a driver of disease,^35–37^ and there are currently no studies which profile the 3D chromatin architecture in human coronary arteries to enable the study of these distal regulatory connections. More recently, technological advances have paved the way for simultaneous multi-omic profiling of the transcriptome and epigenome at single-cell resolution,^32,36,38–45^ presenting a unique opportunity to link genetic variants to cell types and genes to nominate causal gene programs driving disease risk.

Herein, we performed Multiome (RNA + ATAC) sequencing in 44 human coronary arteries to build a multi-omic map of CAD. We utilized novel computational methods^46,47^ to link enhancers to putative target genes and built a single cell variant-to-enhancer-to-gene (V2E2G) map of human CAD. Furthermore, we performed single cell chromatin accessibility quantitative trait loci (caQTL) discovery to prioritize variants casually linked to changes in chromatin. Collectively, we found that the smooth muscle cells (SMCs) harbor the greatest genetic risk of CAD. To expand insight into gene regulation in functionally heterogeneous SMC cell states we utilized dynamic caQTL modeling^48^ to uncover cell-state dependent pathogenicity of disease variants. To dissect the role of genetic variants in regulating larger scale gene networks, we performed genome-wide Hi-C^49^ in 16 human coronary arteries to link variants to distal genes and validated our findings using cell-specific Hi-C with chromatin immunoprecipitation (HiChIP-seq)^50^ and *in vitro* clustered regularly interspaced short palindromic repeats interference (CRISPRi). Collectively, we provide the first multi-omic map of human CAD and integrate transcriptomic, epigenetic, and chromatin architecture to link disease-associated genetic variants with causal cell types and genes.

## Results

### Multi-omic map of human coronary artery disease

We performed single nucleus Multiome (RNA + ATAC) sequencing in 44 human coronary arteries from non-failing donors and patients with chronic heart failure isolated at the time of orthotopic heart transplantation (**Fig. 1a, Extended Data Fig. 1a, Supplementary Table 1**). After quality control^52^, doublet removal^52^, data integration^53^, and clustering, we recovered 126,804 nuclei from 11 distinct cell types (**Fig. 1b, Extended Data Fig. 1b-d**) present in all patient samples (**Fig. 1c**). To annotate cell types, we used paired RNA information from the nuclei for differential gene expression analysis and used canonical marker genes to annotate cell types (**Fig. 1d, Extended Data Fig. 2a-b, Supplementary Table 2**). We used MACS2 to call peaks from the ATAC-seq data (**Fig. 1e**) and identified 143,743 cell type specific differentially accessible marker peaks (**Fig. 1f**). We found that there was a concordance between RNA and ATAC clustering; however, having gene expression enabled annotation of rarer cell types missed by ATAC-based clustering (**Extended Data Fig. 2c-e**). Using the RNA-based annotation and differentially-accessible peaks across cells, we found cell-type-specific enrichment of transcriptional motifs (**Extended Data Fig. 2f**). Having paired RNA and ATAC information from the same nucleus offers a unique opportunity to construct a comprehensive genome-wide enhancer gene map from tissue. We linked peaks to genes using a new supervised single-cell model called scE2G (Wei-Lin Qiu, Maya Sheth, Robin Andersson, and Jesse Engreitz, in preparation), which predicts enhancer-gene regulatory interactions using features based on chromatin accessibility, distance, and peak-gene correlation across single cells (**Fig. 1g**, **Extended Data Fig. 7a-f, Supplementary Table 13-13,** Methods). We found that the non-immune cells (smooth muscle cells (SMCs), endothelial cells (ECs), and fibroblasts) had the greatest number of shared enhancers relative to myeloid cells (**Fig. 1g**).

**Figure 1.**
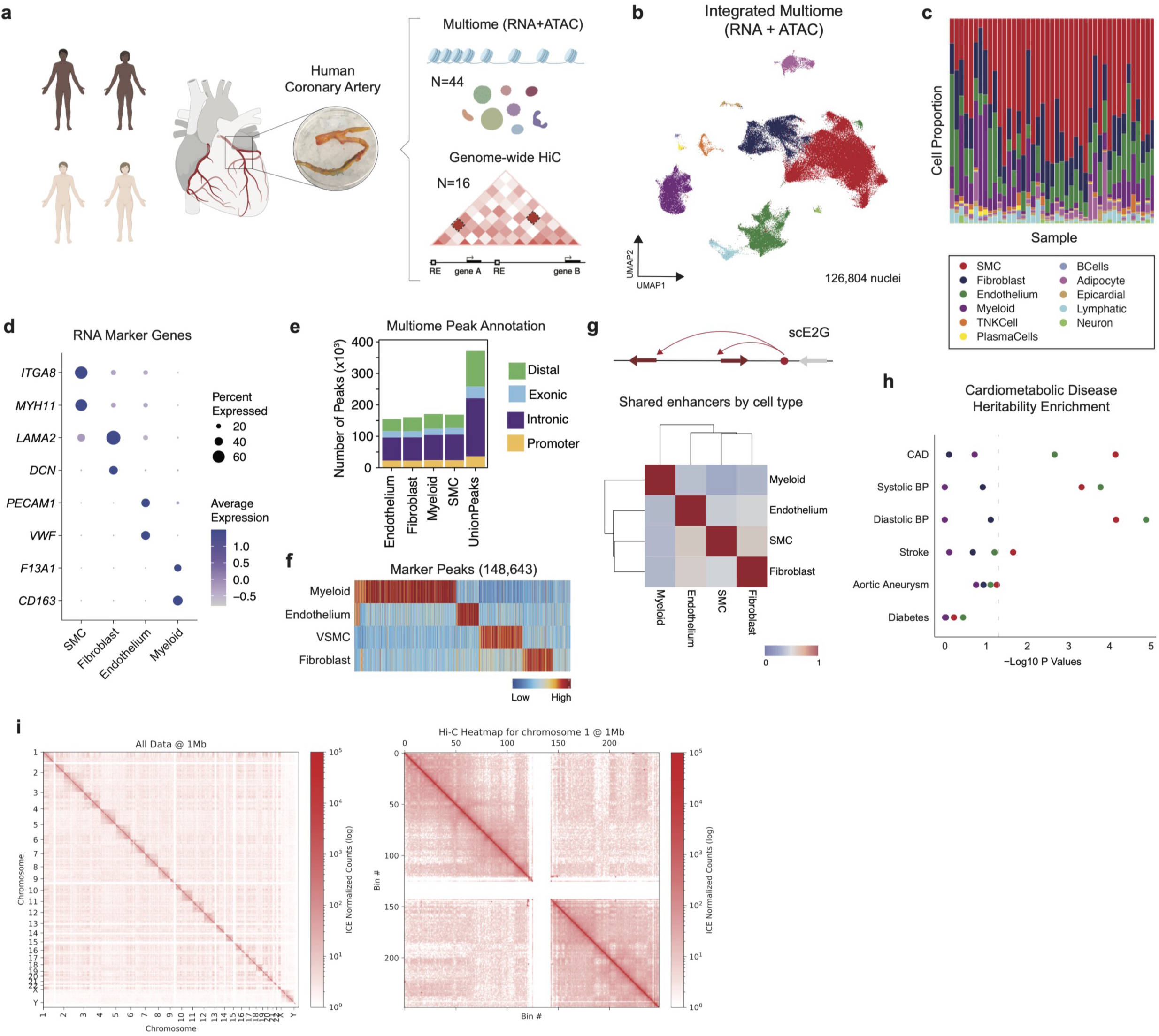
Multiomic profiling of human coronary arteries. (a) Study design. (b) UMAP embedding plot of integrated data clustered by RNA. (c) Cell type composition stack plot by sample. (d) Dotplot of canonical RNA marker genes by cell type; colored by average expression and dot size indicates percent of cells in the cluster which express the gene. (e) Multiome peaks grouped by distal, exonic, intronic, and promoter from macs2 peak calls split by cell type. (f) Differentially accessible marker peaks heatmap by cell type; statistically significant peaks (FDR < 0.1 and log_2_FC > 0.5). (g) Number of shared enhancers heatmap from single cell enhancer-gene map; defined as the number of enhancers where more than 50% of the peak width overlaps with another cell types/number of total enhancers. (h) Linkage disequilibrium score regression for cardiometabolic traits by cell type. (i) HiC heatmap connectivity matrix genome-wide and zoomed in on chromosome 20.

To test for enrichment of disease risk in the four major cell types (SMC, EC, Myeloid, Fibroblast), we used stratified LD score regression^54^ (LDSC) to partition heritability in cell-type marker peaks for key cardiometabolic diseases (coronary artery disease, diabetes, abdominal aortic aneurysms, diastolic/systolic blood pressure, and stroke)^8,55–58^. Notably, we found that SMCs and ECs harbor the greatest genetic risk across cardiometabolic disease variants (**Fig. 1h**). Next, we performed genome-wide Hi-C in 16 human coronary arteries (8 African American and 8 European individuals) (**Fig. 1a,i,j, Extended Data Fig. 3a-d**). We identified more loops at 10 kb resolution than at 2 or 5 kb resolution that were also on average smaller in size (**Extended Data Fig. 3,f**). To further enrich for high-confidence loops, we only retained loops identified in coronaries from at least two individuals. To uncover higher-level chromatin structures that are not as dynamic as loops, we also identified topologically associated domains (TADs) (**Extended Data Fig. 3**).

### Single-nuclei caQTL discovery

Chromatin accessibility quantitative trait loci (caQTL) mapping can identify regulatory variants that regulate accessibility of a regulatory element in the genome through mechanisms such as altered transcription factor binding^51,52^. Single cell caQTLs enable further identification of variants that may regulate chromatin accessibility in a cell-type specific manner^34,53^. To boost our power for single-cell caQTL discovery, we built a chromatin accessibility meta-map by integrating our Multiome ATAC-seq data with prior snATAC-seq from human coronary arteries^34^ (**Extended Data Fig. 4a**). Our integrated CAD snATAC-seq meta-map of genotyped samples included 88 samples and 245,562 nuclei across 4 ancestries (**Extended Data Fig. 4a-c**). We then used this meta-map and RASQUAL^59^ to identify pseudobulk caQTLs in four major cell types (SMC, EC, Myeloid, Fibroblast) (**Fig. 2a, Supplementary Table 14-21**). Notably, RASQUAL simultaneously models allelic imbalance and QTL effect to improve fine-mapping of putative causal variants. A +/-10KB *cis-*window around the peaks was tested for association between variant and chromatin accessibility and age, sex, sequencing site, and the first four principal components of genotype data were included as covariates (see Methods). At the 10% FDR threshold, we found 11,182 caQTLs in SMCs, ECs, Myeloid cells, and Fibroblasts. We found that the power for QTL discovery was associated with number of nuclei (**Fig. 2b, Extended Data Fig. 4d**). As expected, we found that caQTLs were strongly enriched near the peaks they were predicted to regulate (**Extended Data Fig. 4e**). We used the scE2G map to link the caQTL peak to its predicted target gene. We next overlapped our pseudobulk caQTLs with GTEx bulk arterial tissue eQTLs by cell type and found several hundred shared QTL variants in SMCs, ECs, Myeloid cells, and Fibroblasts, respectively (**Fig. 2c, Supplementary Tables 15, 17, 19, 21**). Furthermore, we found a strong correlation in effect size between our SMC caQTLs and bulk coronary artery eQTLs from GTEx (**Fig. 2d**).

**Figure 2.**
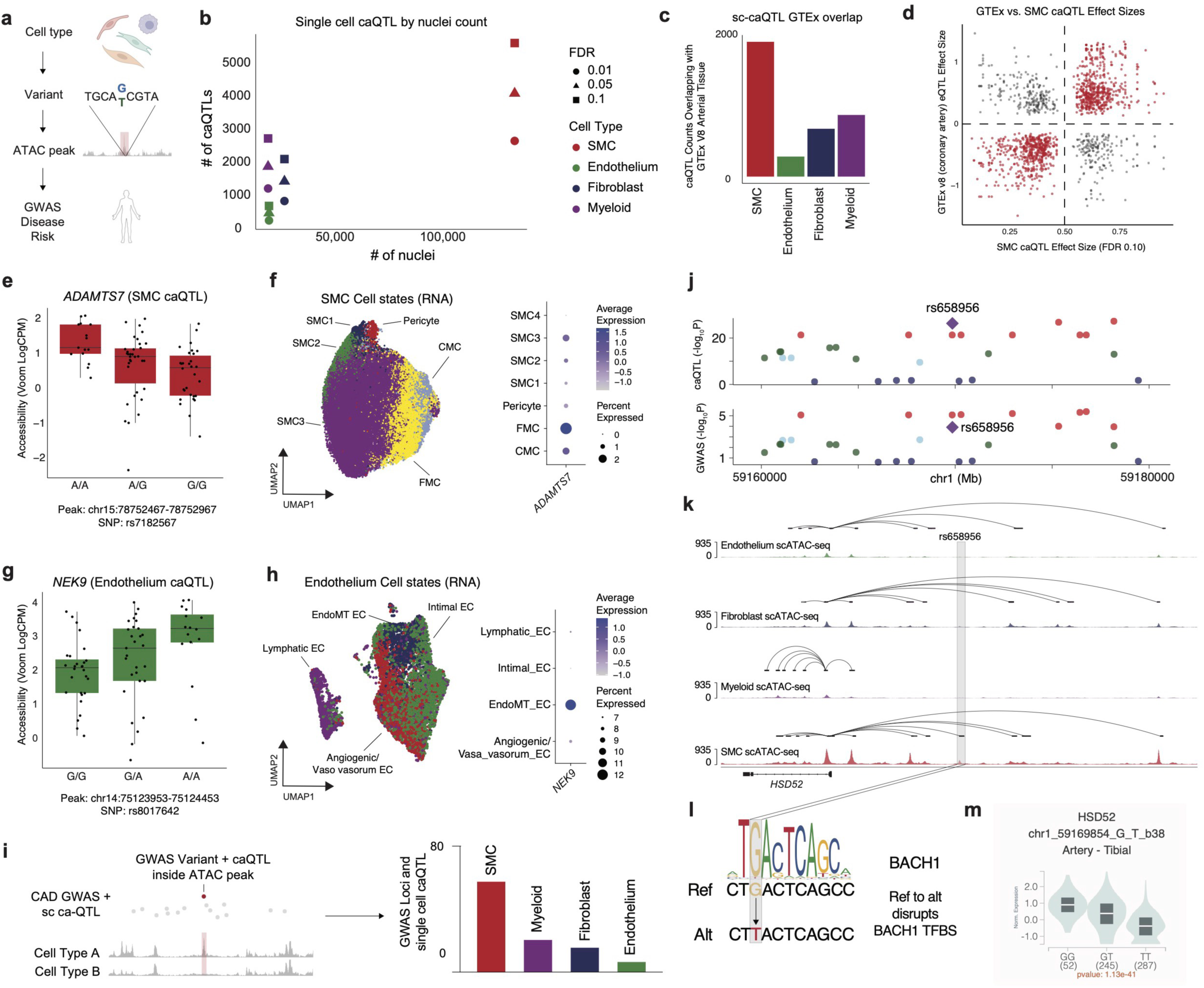
Single-cell chromatin accessibility QT discovery. (a) Schematic of sc caQTL discovery with disease enrichment. (b) Number of pseudobulk caQTL by number of nuclei colored by cell type at various FDR cut-offs. (c) sc caQTL overlap with GTEx arterial eQTL by cell type. (d) GTEx (v8 coronary artery) versus caQTL effect size for FDR < 10% list in SMCs. Rasqual effect sizes are estimated by pi parameter which is centered at 0.5 (less than 0.5 is negative; greater than 0.5 is positive) (e) Box plot of pseudobulk accessibility for peak chr15:78752467-78752967 in SMCs containing disease SNP rs7182567 by genotype. (f) UMAP embedding plot of SMC cell state annotations (left) and dotplot of *ADAMTS7* expression by SMC cell state. (g) Box plot of pseudobulk accessibility for peak chr14:75123953-75124453 in endothelial cells containing disease SNP rs8017642 by genotype. (h) UMAP embedding plot of endothelial cell state annotations (left) and dotplot of NEK expression by endothelial cell state. (i) Number of overlapping sc caQTLs with CAD GWAS loci by cell type. (j) CAD GWAS loci and caQTL Manhattan plot. (k) snATAC-seq tracts with E2G link centered at the rs658956 locus showing SMC specific enrichment of the enhancer with a E2G link to *HSD52*. (l) Ref (G) to Alt (T) at rs658956 disrupts a TF binding site for BACH1 in SMCs. (m) Violin plot showing normalized expression by genotype for rs658956 and *HSD52* in tibial arteries from GTEx bulk data (p-value = 1.1×10^-41^).

To identify disease-associated caQTLs, we overlapped single-cell caQTLs for each cell type with CAD GWAS variants (defined as variants associated with CAD at 1% FDR^8^ along with variants in linkage disequilibrium with these at R^2^ ≥ 0.8 in the European population from 1000 Genomes Project) which yielded numerous disease relevant cell-type-specific caQTLs (**Extended Data Fig. 5a-d**). For example, we found that rs7182567 was an SMC-specific caQTL that was also a GWAS variant (p = 5.4×10^-26^) and a bulk GTEx eQTL for *ADAMTS7* in tibial artery (**Fig. 2e, Extended Data Fig. 5a**). To map this molecular trait to cell state, we leveraged our paired RNA information and sub-clustered the SMCs into transcriptionally distinct cell states, forming a continuum of contractile SMCs (SMC1-3) to de-differentiated SMCs (fibromyocyte, FMC; chondromyocyte, CMC) (**Fig. 2f**). Notably, we found that *ADAMTS7* was most highly expressed in FMCs (**Fig. 2f**) suggesting a role in disease regulation.

Similarly, we found that rs8017642 is an EC-specific caQTL for *NEK9* (**Fig. 2g, Extended Data Fig. 5b**). To identify functionally-distinct endothelium cell states, we mapped our data to a high-resolution single-cell RNA-seq atherosclerosis atlas^60^ and identified four transcriptionally-distinct endothelium subsets (**Fig. 2h**). Notably, *NEK9* was expressed in endothelial cells enriched with endothelium-to-mesenchymal transition (EndoMT) genes, such as *COL1A1* and *FN1*^61^ (**Fig. 2h, Extended Data Fig. 5e**). Pathway enrichment for marker genes for the EndoMT population showed increased EMT and stress response signals (**Extended Data Fig. 5f**). Next, we used the 41 EC-specific putative causal genes previously identified from in a V2G2P analysis of CAD GWAS loci^47^ to create a gene set score which showed maximal enrichment in intimal ECs (**Extended Data Fig. 5g**). Additionally, as prior work has uncovered a gene program^47^ driving CAD pathogenesis in ECs (Program 8), we created a gene signature for all Program 8 genes. Consistent with prior work,^47^ we found an enrichment of this program in angiogenic/vaso vasorum ECs (**Extended Data Fig. 5j**).

To illustrate how a cell type specific caQTL can impart functional transcriptional changes, we examined the CAD GWAS variant rs658956. The variant is an SMC-specific caQTL and regulates a putative enhancer element (chr1:59169569-59170069) that is linked to the *HSD52* gene (based on scE2G) (**Fig. 2j,k**). Additionally, transcription factor binding site (TFBS) analysis found that rs658956, which changes the reference (G) to alternative (T) allele, is predicted to disrupt a BACH1/2 TF binding motif (**Fig. 2l**). Furthermore, we found that rs658956 is also a tibial artery eQTL for *HSD52* expression (p = 1.1×10^-41^) wherein the switch from reference (G) to alternate (T) is associated with decreased expression (**Fig. 2m**). Collectively, this would suggest that the GWAS variant rs658956 may impact TF binding thereby decreasing chromatin accessibility of the enhancer and subsequently reducing *HSD52* gene expression.

### Idenfitying caQTLs that are dynamic with SMC de-differentiation state

Molecular QTL discovery using bulk or pseudobulk data is generally carried out in a cell state agnostic-manner from tissues which collapses cells across a heterogeneous mix of cell states. For example, in the development of atherosclerosis is it known that SMCs undergo a phenotypic switch from a contractile to a fibroblast-like FMC phenotype^30^ which is accompanied by corresponding transcriptional changes. Paired multiomic RNA-ATAC sequencing offers the opportunity to characterize individual cells within a global cell type across continuous transcriptional cell states and to model this continuum of heterogeneous cell states to discover peaks that may modify or be modified by QTL variant effects^54^.

Using the top 100 marker genes defined in FMCs from Wirka *et al*^30^, we created an FMC module score across our integrated SMC map and identified a continuum of activation states (see Methods, **Fig. 3a, Extended Data Fig. 6a-b**). We then binned nuclei into bottom, middle, and top thirds based on FMC score for downstream modeling (**Fig. 3b**). To dissect caQTL dynamics in the context of cell state, we utilized a Poisson mixed effect (PME) single cell model^48,55^ (**Fig. 3c, Extended Data Fig. 6c**). This approach models cell-level fragment counts with random effect covariates to account for relatedness shared by cells from the same sample and sequencing conditions. Briefly, at baseline we model ATAC fragment counts with fixed effect covariates including genotype, number of fragments per cell, TSS enrichment per cell, principal components of single-cell ATAC counts, principal components of genotype data, random effect covariates (batch and sequencing site), and intercept. To test for dynamic caQTLs where the genotype effect is modified by SMC cell state, we then added a cell-level FMC score term and a genotype-FMC score interaction term as covariates in the model (**Fig. 3c**). Finally, we binned the nuclei by bottom, middle, and top FMC score buckets (**Fig. 3b,c**) and identified dynamic caQTLs which differ according to cell-state.

**Figure 3.**
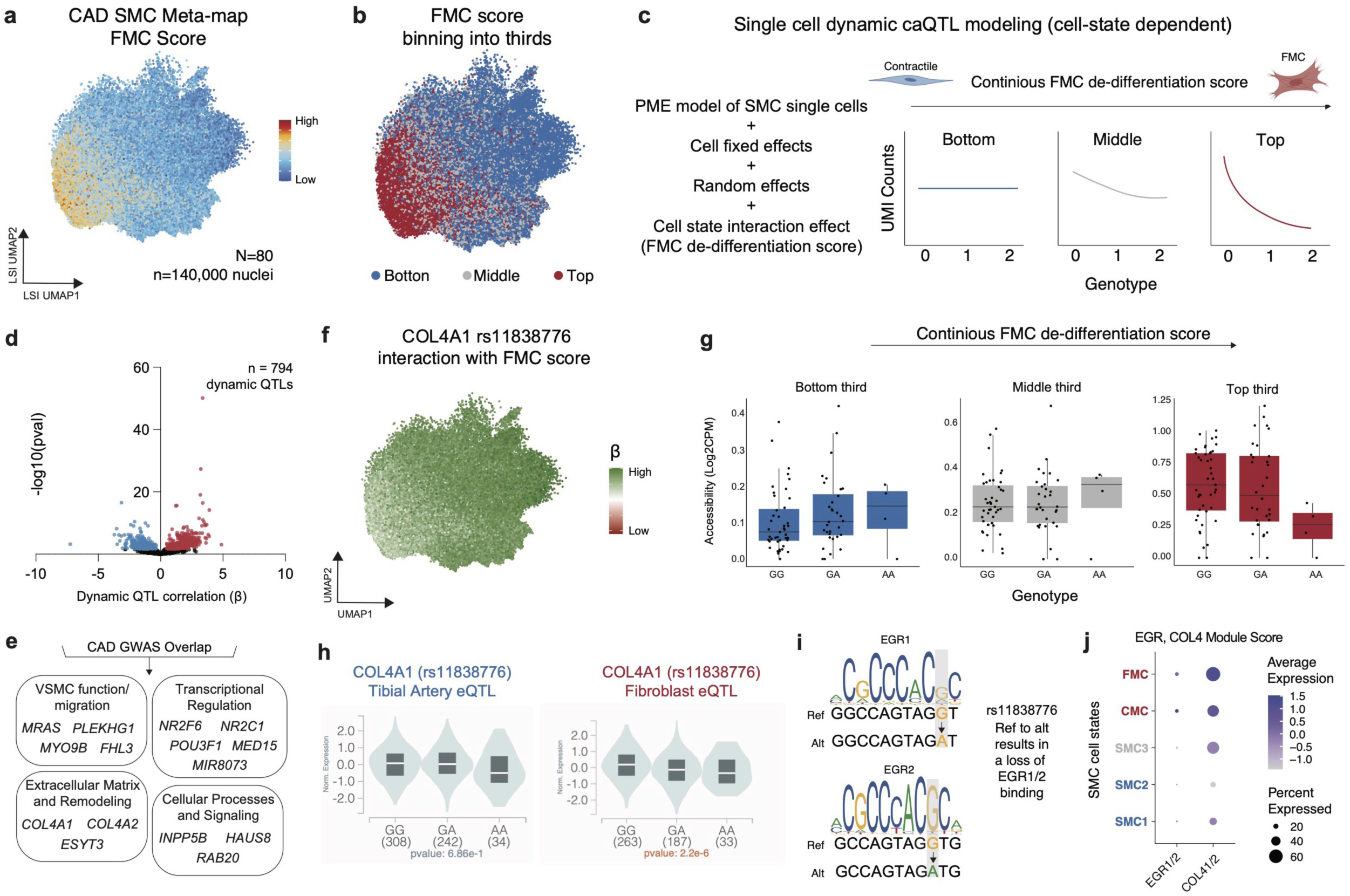
Dynamic QTL discovery in SMC de-differentiation in atherosclerosis. (a) FMC gene set score (top 100 genes from Wirka *et al*^30^ for FMC marker gene) in integrated SMC meta-map. (b) Nuclei binned into tertiles based on FMC score. (c) Dynamic single cell caQTL modeling strategy using the PME model for cell-state dependent single nuclei caQTL analysis. Covariate adjustment and interaction between a continuous FMC cell state score (binned into low, medium and high) and genotype (see Methods). (d) SMC single cell dynamic caQLTs volcano plot showing - log10(adjusted p-vaue) by interaction coefficient (*β*_total_ = *β*_G_ + FMC score × *β*_FMC_); dots are colored by: black = not significant, blue/red = adjusted p-value < 0.05, where blue and red represent *β*_total_ < 0 and *β*_total_ > 0 respectively. (e) Genes linked to dynamic caQTLs which are CAD GWAS loci using E2G grouped by biological category using ChatGPT. (f) Interaction coefficient of the rs11838776 caQTL for *COL4A1* in SMCs with FMC score. (g) Box plots show the caQTL effect for SMCs in the bottom (left), middle (center), and top (right) thirds of FMC scores. (h) rs11838776 eQTL in tibial artery (left) and cultured fibroblasts (right) for *COL4A1* showing no correlation in tibial arteries but a negative correlation in fibroblasts by genotype. (i) rs11838776 ref (G) to alt (A) removes a EGR1/2 binding site. (j) *EGR1 + 2* and *COL4A1 + 2* gene set score dotplot by SMC cell state showing greatest expression in CMC/FMC cell states.

We ran the PME single cell model on the lead variants of SMC-specific caQTLs identified through the RASQUAL pseudobulk analysis and found 794 caQTLs which are dynamic with respect to FMC score (**Fig. 3d, Supplementary Table 22**). The significant interaction term indicates a modification of genotype effect on peak accessibility during cell-state transition or a genotype-specific modification of cell-state dependent changes on peak accessibility, both of which are plausible depending on the mechanism of the variant (e.g. variant modifies pioneer vs. non-pioneer transcription factor binding). Notably, of the 794 ATAC peaks regulated by dynamic caQTLs, 12 also contained CAD GWAS variants (**Fig. 3e**). We then used the scE2G map to link the 12 dynamic disease-associated caQTLs to target genes. To connect the dynamic disease-associated genes to gene programs, we used a large language model (ChatGPT) to group genes by biological groups and found an enrichment of VSMC function/migration, transcriptional regulation, and ECM remodeling cellular processes and signaling pathways (**Fig. 3e**). Interestingly, we found that a gene set score of these genes (**Fig. 3e**) was enriched in FMCs (**Extended Data Fig. 6d**) bolstering a role in disease regulation through cell state phenotypic switching. For example, the CAD GWAS SNP rs11838776 is a dynamic caQTL variant for an enhancer (chr13:110388102-110388602) that lies within *COL4A2/COL4A1* locus. The strongly negative interaction term of the PME model suggests that the alternate allele of rs11838776 may further decrease accessibility of this enhancer as the SMCs transition into a more FMC-like cell state. We further confirmed this effect by visualizing minimal genotype-specific effect in ATAC accessibility in “quiescent” SMCs (bottom third FMC score) versus larger negative genotype-specific effect in “de-differentiated” FMCs (top third FMC score) (**Fig. 3f,g**). Interestingly, the bulk GTEx data suggests that rs11838776 is not an eQTL for *COL4A1* in tibial arteries (p = 0.69) but is an eQTL for *COL4A1* in cultured fibroblasts (p = 2.2×10^-6^) with the same directionality as the interaction effect (**Fig. 3h**). Collectively, this would suggest that rs11838776 and its enhancer may regulate *COL4A1* expression only as cells enter a fibrogenic fate. To dissect the molecular mechanism driving this dynamic QTL effect, we explored TF binding motifs surrounding the rs11838776 variant. We found that the reference allele (G) contributes to a consensus binding motif for EGR1/2, while the alternate allele (A) is predicted to abrogate this motif (**Fig. 3i**). Furthermore, we created a gene set score for *EGR1/2* and *COL4A1/2*, respectively, using RNA data and found greatest expression in the CMC/FMC cell state suggesting that as SMCs de-differentiate *EGR1/2* expression increases, driving *COL4A1/2* expression (**Fig. 3j**). We also found that the CAD GWAS variant rs658956 was dynamic for chromatin accessibility at the SMC specific enhancer and linked to *HSD52* (**Fig. 2j-k, Extended Data Fig. 6e**).

### Single cell disease variant-to-enhancer-to-gene map

To map CAD disease variants to cell types and genes in an unbiased fashion, we built a disease relevant variant-to-enhancer-to-gene (V2E2G) map as follows^46,47,63^ (**Fig. 4a,b**):

1. Select ATAC-seq peaks from the 4 major cell types which contain a CAD GWAS variant (1,533)
2. Variant-to-enhancer (V2E): identify cell-type-specific peaks (as called by MACS2, see Methods) overlapping disease variants, and H3K27ac peaks from bulk coronary arteries^56,57^. Using this approach, we found cell specific V2E pairs: SMC (490), endothelium (424), fibroblast (431), and myeloid (367).
3. Variant-to-enhancer-to-gene (V2E2G): Link peaks from (2) to genes using a new supervised single-cell model called scE2G (Wei-Lin Qiu, Maya Sheth, Robin Andersson, and Jesse Engreitz, in preparation), which predicts enhancer-gene regulatory interactions using features based on chromatin accessibility, distance, and peak-gene correlation across single cells (**Fig. 4a, Extended Data Fig. 7a-f**, see Methods). Overlapping scE2G predictions with our cell-type-specific peaks, we found the following (single-cell V2E2G peaks, no. linked genes): SMC (276, 325), endothelium (239, 268), fibroblast (235, 279), and myeloid (199, 236).
4. Finally, to nominate V2E2G links which also affect a molecular trait, we overlapped V2E2G peaks with our single cell caQTLs and found the following (single-cell V2E2G peaks with caQTLs, no. linked genes): SMC (49, 41), endothelium (5, 3), fibroblast (13, 11), and myeloid (15, 12).

**Figure 4.**
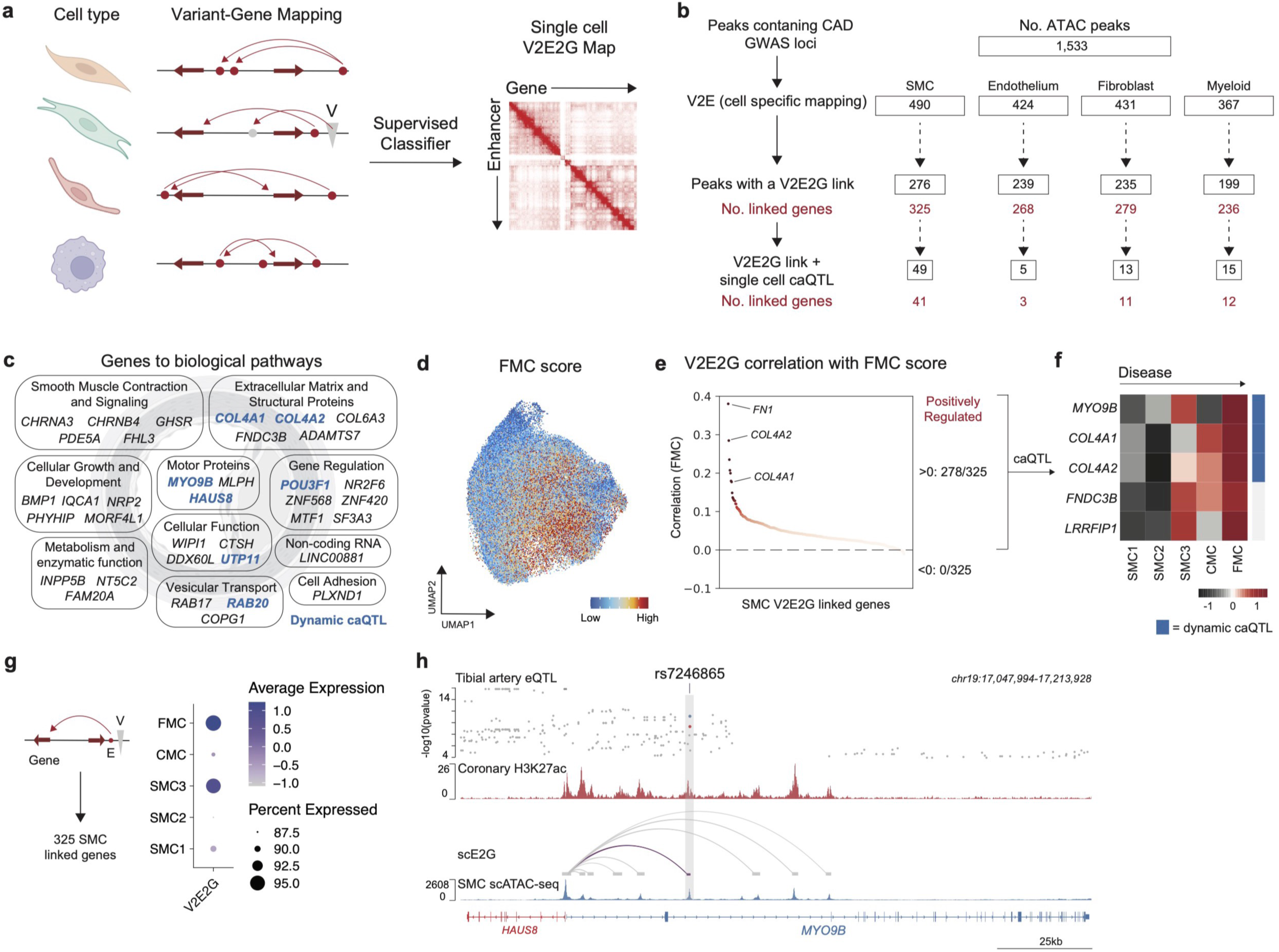
Building a single cell variant to enhancer to gene (scV2E2G) map of coronary artery disease. (a) Model schematic for single cell V2E2G mapping using a supervised classifier. (b) Number of CAD GWAS variants mapping to snATAC-peaks and linked genes across cell types with overlapping caQTLs. (c) 41 SMC scV2E2G and caQTL grouped by biological category using ChatGPT; genes highlighted in blue are also genes linked to a dynamic caQTL peak. (d) FMC gene set score (top 100 genes from Wirka *et al*^30^ for FMC marker gene) in SMC UMAP embedding plot. (e) Correlation between FMC score and combined gene-set score for 325 genes identified from V2E2G map in SMCs colored by p-value. (f) Heatmap for SMC V2E2G links positively correlated with FMC which are also single cell caQTLs by grouped by SMC cell state showing increasing expression in diseased SMC cell states; blue indicates genes linked to a dynamic caQTL peak. (g) Gene Set score for the 325 genes identified from V2E2G map in SMCs grouped by SMC cell state. (h) ENCODE coronary artery H3K27ac and snATAC-seq SMC tract at rs7246865 SNP with scE2G link to *MYO9B* and *HAUS8*.

Notably, we found that SMCs were most enriched with V2E2G links and likely harbor the greatest genetic risk of driving CAD. Overlapping V2E2G with single-cell caQTLs provides a high-fidelity set of cell-type-specific hits which may be driving disease in SMCs through a direct change in a molecular trait – however, this approach is challenging in more rare cell types where fewer nuclei hinder QTL discovery (**Fig. 2b, 4b**).

### SMC V2E2Gs are disease relevant

We utilized ChatGPT to group SMC V2E2G which are caQTLs (41 genes) into functional biological programs and found enrichment across SMC processes including ECM remodeling, cellular growth and proliferation, metabolism, cytoskeletal organization, and gene regulation (**Fig. 4c**). To identify SMC V2E2G targets which correlate with disease burden, we first used the top 100 FMC marker genes from Wirka *et al*^30^ to generate a FMC marker gene score (**Fig. 4d**), correlated expression of genes identified from V2E2G in SMCs with FMC score, and plotted the correlation colored by adjusted p-value (**Fig. 4e**). We found that 278/325 genes from our V2E2G showed a positive correlation with FMC score while no genes had a negative correlation, demonstrating an enrichment for disease relevant linked genes (**Fig. 4e**). To identify those with highest confidence, we restricted these 278/325 genes to those that were nominated from our bulk arterial GTEx and SMC caQTL overlap analysis which resulted in a set of 5 genes (**Fig. 4e,f**). A heatmap of these 5 genes (*MYO9B, COL4A1, COL4A2, FNDC3B,* and *LRRFIP1*) shows increasing expression as SMCs transition from contractile (SMC1-3) towards de-differentiated disease associated states (CMC, FMC) (**Fig. 4f**). Furthermore, we created a gene set score for the 325 SMC V2E2Gs and found greatest enrichment in SMC3 and FMCs (**Fig. 4g**) – collectively, these findings support the notion that FMCs harbor the greatest genetic risk of CAD. Among these, we highlight rs7246865 as an example as we found this variant to be an SMC dynamic caQTL, a pseudobulk caQTL, and a V2E2G link, thus underscoring its potential importance in disease. We show that rs7246865 falls inside an SMC peak (snATAC-seq) and coronary artery enhancer (ENCODE human coronary artery H3K27ac), and there is a E2G link to the *MYO9B and HAUS8* transcription start site (**Fig. 4h**). Importantly, a prior study has used CRISPRi to validate that disruption of rs7246865 is causally linked to *MYO9B/HAUS8* expression^8^.

### Disease variant chromatin networks

To dissect the broader impact of disease variants on gene networks through chromatin looping, we used genome-wide human coronary HiC to construct large scale chromatin networks (**Fig. 5a**). Briefly, we built interconnected chromatin networks with interacting anchors using a fast greedy modularity optimization algorithm^58,59^(**Fig. 5a**, see Methods). We found 5,331 interconnected networks (**Fig. 5b**). To identify disease associated networks and build a variant-to-enhancer-to-network (V2E2N) map, we identified networks where any loop within the network contains a V2E in either anchor (i.e. a snATAC peak that contains a GWAS variant and overlaps with and ENCODE coronary artery H3K27ac) for SMCs, fibroblasts, ECs, and myeloid cells (**Fig. 5b**). Interestingly, we found that all networks contain an average of 18 HiC loops while disease associated networks (V2E2N) contain an average of 59 HiC loops (in SMCs) (**Fig. 5c**). To identify genes implicated in our V2E2N, we found all snATAC-seq peaks present in any HiC anchors in the V2E2N (**Fig. 5a**) and identified linked genes using the scE2G map as before (**Fig. 4a,b**). Using this interested approach, we mapped a variant to a catalogue of genes that may be regulated locally or distally through a linked chromatin network (**Fig. 5d**). This strategy identified key disease networks which form ‘super chromatin interactome’ hubs wherein a disease variant is part of several hundred interconnected chromatin loops (**Fig. 5d,e**). Interestingly, we found that the CAD GWAS variant rs4887091 (located within enhancer element chr15:78750919-78751419) was part of a super chromatin interactome that consisted of numerous genes such as *CHRNA3, CHRNA4, MORF4L1, CHRNA5, CTHS,* and *ADAMTS7* which were also V2E2G for rs4887091 – notably, *CHRNA3, CHRNA4,* and *ADAMTS7* were also arterial eQTLs for rs4887091 (**Fig. 5e,f**). At the disease variant rs4887091, a base pair change from the reference (T) to alternate (C) allele leads to a change in a CTCF binding motif (**Fig. 5g**). We performed ChIP-seq for CTCF in cultured human coronary artery smooth muscle cells (HCASMCs) and demonstrated that rs4887091 overlaps a CTCF binding site, suggesting that this variant may drive broader changes in chromatin organization through regulation of CTCF binding (**Fig. 5h**).

**Figure 5.**
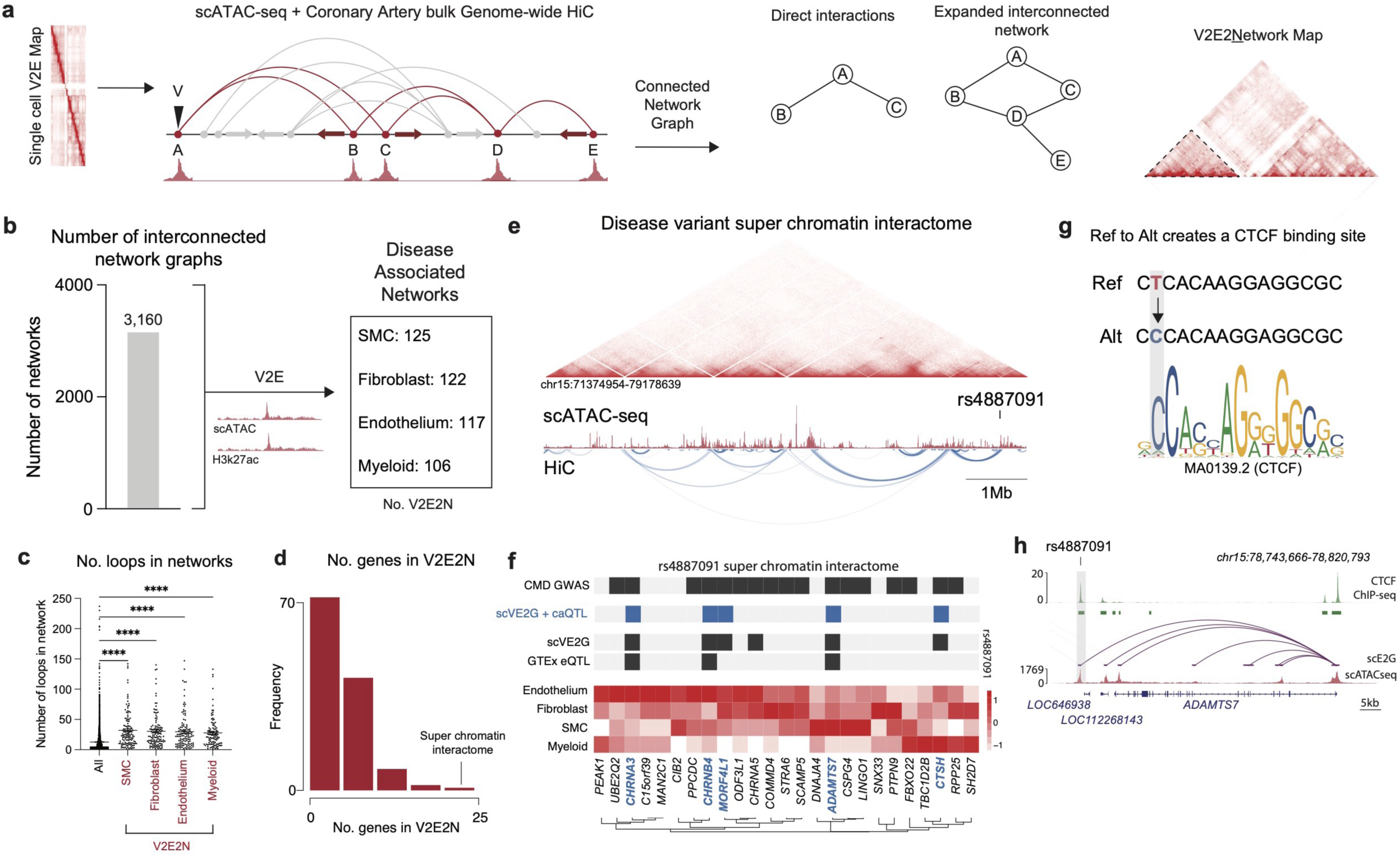
Building a variant to enhancer to network (scV2E2G) map of coronary artery disease. (a) Analysis framework for constructing interconnected graph networks using scV2E and genome wide tissue HiC. (b) Number of distinct network graphs identified from HiC loop connectivity (left) and filtered for networks which contain a cell specific V2E. (c) Number of loops in all networks and disease enriched networks (networks where any HiC anchor contains a CAD GWAS variant inside a coronary artery enhancer) by cell type. (d) SMC V2E2N: number of linked genes in networks which contain a disease variant (V2E) in SMCs. (e) Disease variant rs4887091 super chromatin interactome showing SMC snATAC-seq and coronary HiC heatmap with looping between chr15:71374954-79178639. (f) Genes in the rs4887091 super chromatin interactome; heatmap showing expression by cell type, caQTL/GTEx eQTL/scE2G for rs4887091, and CMD GWAS related genes. (g) rs4887091 ref (T) to alt leads to increased CTCF binding. (h) CTCF ChIP-seq in HCASMCs overlaid with SMC snATAC-seq and scE2G at the rs4887091 locus.

### Cell type specific mapping of disease variants to distal genes

Although our scE2G predictions are limited to enhancer-gene interactions within a *cis-*window, leveraging our Hi-C based V2E2N analyses we can assign potential distal gene regulatory function to disease variants. Using our V2E2N, we can map disease variants to distal genes. As an example, we find that the CAD GWAS variant rs9876658 falls within a snATAC-seq peak in SMCs, ECs, and fibroblasts (but not myeloid cells) highlighting stroma specific regulation (**Fig. 6a**). From the genome-wide coronary artery HiC we find a loop connecting the rs9876658 enhancer to the *AMOTL2* transcription start site – notably, we do not find a chromatin loop between rs9876658 and the nearest gene *ANAPC13* (**Fig.6a**). Collectively, this analysis predicts that rs9876658 regulates *AMOTL2* expression in stromal cells, however since our HiC is from bulk coronary arteries we cannot implicate a specific cell type, and furthermore, snATAC-seq alone does not identify functional enhancers.

**Figure 6.**
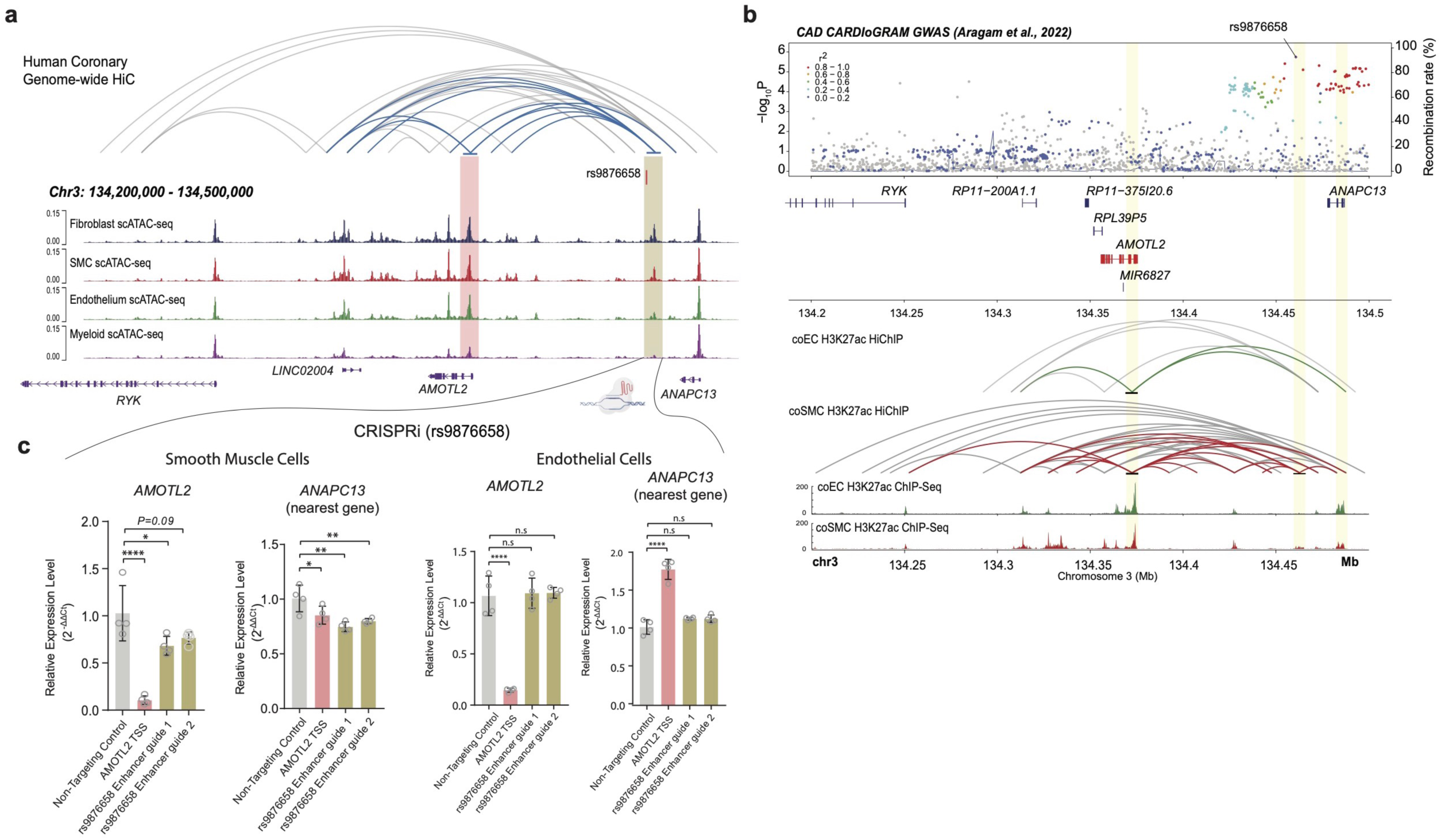
rs9876658 distally linked to *AMOTL2* in SMCs. (a) snATAC-seq tracts and genome wide coronary HiC loops showing rs9876658 lies within a non-immune peak linked distally to the *AMOTL2* transcription start site. (b) Manhattan plot for rs9876658 overlaid on HiChIP-seq and H3K27ac from primary SMC and endothelial cells showing rs9876658 lies within a SMC specific enhancer and is linked to the *AMOTL2* TSS in SMCs but not endothelial cells. (c) CRISPRi for enhancer containing the rs9876658 variant in SMCs and endothelial cells shows that rs9876658 regulates expression of *AMOTL2* and ANAPC13 in SMCs but not endothelial cells. CRISPRi for a non-targeting (negative control) and TSS (positive control) shown as reference.

Furthermore, published bulk coronary artery H3K27ac does not provide cell specificity for active enhancers, so to map disease variants to cell type specific enhancers, we performed H3K27ac ChIP-seq in human coronary artery smooth muscle cells (HCASMC) and human coronary endothelial cells (HCAEC) and found that rs9876658 overlapped with a H3K27ac peak in SMCs but not ECs (**Fig. 6b**), suggesting the ATAC-seq peak containing rs9876658 is a functional enhancer in SMCs but not endothelial cells. Next, to address limitations of bulk tissue chromatin capture, we performed HiChIP-seq in HCASMCs and HCAECs and found a direct loop between the enhancer containing rs9876658 and the *AMOTL2* promoter in SMCs but not ECs (**Fig. 6b**).

### CRISPRi of rs9876658 affects *AMOTL2* expression in SMCs

To causally link rs9876658 to cell specific regulation of *AMOTL2*, we performed CRISPRi targeting the TSS (*AMOTL2*) and regulatory enhancer (containing rs9876658) with a Lenti-dCas9-KRAB-blast vector (see Methods). We found that targeting the *AMOTL2* TSS led to a complete knockdown of *AMOTL2* expression in HCASMCs and HCAECs validating our system (**Fig. 6c**). Interestingly, targeting the *AMOTL2* TSS also led to a modest reduction in *ANAPC13* expression in HCASMCs, consistent with our network analysis which identified *AMOTL2* and *ANAPC13* being part of the same network (**Fig. 6c**). Importantly, targeting of the enhancer containing rs9876658 led to a reduction in *AMOTL2* expression in HCASMCs but not HCAECs bolstering our prediction from human tissue mapping (**Fig. 6c**). Collectively, these analyses nominate rs9876658 as a causal regulator of *AMOTL2* expression in SMCs but not ECs and implicate this gene as a novel candidate for atherosclerosis.

## Discussion

GWAS studies have identified hundreds of variants driving common diseases such as CAD^1,7–9,60^. A key challenge in going from human genetics to druggable therapies is our lack of understanding of the causal cell types and gene programs driving disease^15^. The emergence of multi-omic single cell technologies has given us a unique opportunity to profile human tissue at high resolution^25,61^. Numerous studies have leveraged these technologies to uncover cellular heterogeneity in human tissue across many diseases contributing to a human cell atlas^26–29,32,33,62,63^. Yet, there are few studies which have mapped common disease variants to causal cell types and genes for target discovery from human tissue. Further, a key limitation of prior studies is the use of uni-modal sequencing technologies or reliance on cell-based systems which precludes discovery of disease targets from the native tissue environment.

Our study utilized a new technique, single nucleus Multiome (RNA + ATAC)^32,38,39^, in human coronary arteries from 44 patients with paired genotyping for un-biased characterization of the transcriptome and epigenome at single cell resolution. Capturing paired gene expression and chromatin accessibility from the same nucleus allows for ground truth peak-gene linkage^40,42,43,64,65^ and is an important milestone for gene regulation discovery in human tissue. We also utilized a novel computational framework, scE2G^46,47,66^, to build an un-biased genome-wide single cell V2E2G map in human CAD. Notably, we found that SMCs were most enriched with V2E2G links (325 linked genes) and likely harbor the greatest genetic risk of driving CAD. This systematic approach can be applied across diseases and allows for an unbiased framework to prioritize pathological genes and cell types driving human disease.

To validate our scV2E2G map using an orthogonal strategy in a broader patient cohort, we integrated our data with published human coronary artery snATAC-seq^34^ to build a meta-map of 88 samples for single cell caQTL discovery. Using this strategy, we discovered 11,182 single cell caQTLs many of which overlapped with bulk arterial tissue GTEx eQTLs. Notably, this approach allows for discovery of novel QTLs in rarer cell types that will be missed by traditional bulk approaches^54,67^. To enrich for disease associated QTLs, we overlapped our scV2E2G map with caQTLs and to nominate a high confidence gene set with GWAS, scE2G, and molecular trait evidence in regulating disease – we found 5 such genes (*MYO9B, COL4A1, COL4A2, FNDC3B,* and *LRRFIP1*) whose expression was also correlated with the SMC fibromyocyte transition state.

Traditional pseudobulk QTL discovery is a cell state agnostic method^34,68^ and lacks the ability to discriminate between molecular traits which are influenced by disease state^48^. Here, we utilize single cell genomics in a large population to incorporate cell state dynamics in one of the most fluid cell states in the atherosclerotic plaque – SMCs. In atherosclerosis, contractile SMCs undergo de-differentiation and acquire a fibromyocyte phenotype (FMC) with distinct transcriptional signatures^30,69,70^. We performed single cell caQTL discovery using the FMC de-differentiation score as a continuous variable to identity QTLs which are cell state dependent. Using this framework, we uncovered rs11838776 as a QTL for *COL4A1* which dynamically becomes active as SMCs de-differentiate via a TF binding site change for EGR1/2. Interestingly, *EGR1*/2 are universal stripe factors which are known to orchestrate changes in chromatin accessibility for recruitment of co-binding factors^71,72^. This is the first application of single cell state dependent QTL modeling in human tissue and can expand our understanding of gene regulation in heterogenous cells from tissue. In cases where there are epigenetically or transcriptional defined cell states and cell state trajectories that may be disease relevant, such approaches present an efficient way to uncover molecular QTLs that may modify or be modified by disease-associated differentiation process that may be missed while profiling a bulk tissue.

Given the genome is a dynamic 3D structure^73^, chromatin looping through TADs is a crucial high-level state governing downstream gene regulation^73–76^. Currently, there are no studies which profile chromatin looping in human CAD. Prior studies in cell-based systems and other tissues have demonstrated the power of HiC technologies to uncover chromatin architecture in the context of gene regulation and mapped disease variants to distal causal genes^77–86^. Here, we performed genome-wide HiC in 16 human coronary arteries and used network graphs to connect chromatin communities. This approach allows for building tissue-specific gene regulatory networks within TADs. Numerous studies have shown that large scale chromatin structures have tissue and cell specificity^78,80,87^, yet no study has profiled looping in diseased human arterial tissue. We integrated our chromatin networks with the scV2E2G map to identify disease variants enriched in a super chromatin interactome that may regulate a broader gene program. To our surprise, we found that the disease variant rs4887091 drives a super chromatin interactome through a modified CTCF binding site. Given the role of CTCF in regulating TAD insulation boundaries, these results would suggest that this disease variant is orchestrating broad scale chromatin changes. Furthermore, by integrating HiC tissue looping with multi-omic scV2E2G mapping, we uncovered several novel candidate genes linked to CAD GWAS loci that were not previously implicated. We found rs9876658 was linked to the distal candidate gene *AMOTL2* and used cell specific H3K27ac and HiChIP-seq and CRISPRi to target the active enhancer containing rs9876658 to show SMC specific regulation.

Our study is not without limitations. First, we rely on nuclear RNA from multi-omic capture which precludes discovery or cellular transcripts as prior studies have shown that single cell RNA is deeper for cell state annotation. We utilized high resolution published single cell data to perform reference mapping for state imputation^88^. Second, QTL associations can have pleotropic effects making it challenging to parse out relevant targets from our large caQTL discovery analysis – we leveraged cell specific QTL discovery and single cell caQTL modeling to enrich for and prioritize disease associated cell types and cell states. Third, our coronary artery HiC data is bulk precluding inference of cell type specific looping – to prioritize cell types and increase confidence for cell specific looping, we used HiChIP-seq in targeted cell types in vitro.

In conclusion, we built the first multi-omic gene expression and chromatin accessibility map of human CAD and provide a comprehensive framework to map CAD GWAS variants to cell types and genes. Additionally, we use single cell QTL modeling to characterize state dependent pathogenicity of disease variants in human tissue. We used tissue HiC to build large scale chromatin networks and uncover how disease variants impact distal gene regulation. Finally, we integrated tissue omics with cell based epigenetic profiling to prioritize and functionally test candidate genes using enhancer TAP-seq and CRISPRi. Collectively we provide a disease agnostic framework to translate human genetic findings to identify pathologic cell states and genes driving disease – this study provides a comprehensive V2E2G map with genetic and tissue level convergence for future mechanistic and therapeutic studies.

## Acknowledgments

JMA was supported by the American Heart Association Predoctoral Fellowship (826325) and is currently supported by Leducq Foundation Network Seed Grant (#20CVD02). JMA and PL are supported by the Washington University School of Medicine Medical Scientist Training Program. This work was supported by National Institutes of Health (NIH) grants UM1HG008853 (IMH and NOS), R01HG013371 (IMH and NOS), R01HL159171 (NOS), R01HL171045 (TQ), R01HL134817 (TQ), R01HL139478 (TQ), R01HL156846 (TQ), R01HL151535 (TQ), R01HL158525 (TQ), UM1HG011972 (JME and TQ), U01HG011762 (TQ), R01HL159176 (JME), R01HL164811 (RG and JME), American Heart Association 23SCISA1144703 (PC), 24SCEFIA1248386 (PC), 695 20CDA35310303 (PC), and the Novo Nordisk Foundation Center for Genomic Mechanisms of Disease (NNF21SA0072102). MUS acknowledges the support of an NSF Graduate Research Fellowship (DGE-1656518) and a graduate fellowship award from Knight-Hennessy Scholars at Stanford University. NOS was also supported in part by the Foundation for Barnes-Jewish Hospital. We thank Dr Matthew-Ackers Johnson from NUS Cardiovascular Research Institute for the provision of the primary HCAEC and HCASMC cell lines. RSYF is funded by Individual Research Grants from the National Medical Research Council (NMRC) of Singapore (MOH-001480-00) and MOE Academic Research Fund (AcRF) Tier 3 (MOE-000333-00). The study was partially supported from an Amgen sponsored research agreement. Study design schematics were created in BioRender.com. We thank the Genome Technology Access Center at the McDonnell Genome Institute at Washington University School of Medicine for help with genomic analysis. The Center is partially supported by NCI Cancer Center Support Grant #P30 CA91842 to the Siteman Cancer Center. This publication is solely the responsibility of the authors and does not necessarily represent the official view of the NIH.

## Author Contributions

NOS and IMH conceived the study. JMA, TQ, and NOS drafted the manuscript with assistance from all authors. JMA collected all coronary arteries. JMA and AB isolated nuclei from human coronary arteries. JMA performed 10x Multiome cDNA construction for library preparations and sequencing. JMA, PCL, IE, CJK, MS, WLQ, SK, DYL, DL, AT, PC, and QZ performed all computational analysis. CLM assisted with QTL discovery. TY, KL, SJ, BA, CML, and YHH assisted with the HiC study. CJML, RG, and RSYF performed all CRISPRi experiments. CAN, QZ, MR, and CJML performed all in vitro HiChIP and H3K27ac experiments. RG and JME assisted with EC specific analyses. REW and AA assisted with in vitro experiments. MUS, WQ, RA, and JME developed and applied scE2G. TQ, RG, RSYF, SH, and SJ provided guidance on GWAS mapping and data interpretation. All authors contributed to the experimental design, data analysis and interpretation as well as manuscript production. NOS is responsible for all aspects of this manuscript including experimental design, data analysis, and manuscript production. All authors approved the final version of the manuscript.

## Materials and Methods

### Ethical Approval for Human Specimens

The study is compliant with all relevant ethical regulations and has been approved by the Washington University School of Medicine Institutional Review Board (IRB #201104172). Informed consent was obtained from each patient prior to tissue collection by Washington University School of Medicine and no compensation was provided in exchange for subject participation in the study. All demographic and clinical data has been de-identified and provided in Supplementary Table 1. Patients included in this study span diverse race, age, and sex to provide an inclusive trans-ethic study population.

### Inclusion Criteria

Prior to tissue collection, specific inclusion criteria were employed to ensure well controlled study groups. Any patients with HIV or hepatitis and known genetic cardiomyopathies were excluded from this study. The left anterior descending coronary artery was isolated, and flash frozen from donor hearts: patients with stable ejection fractions, no known history of cardiac disease and experienced a non-cardiac cause of death/transplant and from patients with chronic heart failure. For all samples the proximal left anterior descending coronary artery was used.

### Nuclei isolation for Multiome sequencing

The left anterior descending coronary artery was dissected from explanted hearts, epicardial fat removed, and arteries were flash frozen using liquid nitrogen. Identical regions from the proximal left anterior descending artery were used from all patients. Single nuclei suspensions were generated as previously described^32^. Nuclei were isolated according to 10x Genomics protocol (CG00375; Nuclei Isolation Complex Sample for ATAC GEX Sequencing RevB) and flow cytometry for 7-AAD (Sigma; SML1633-1ML) positive nuclei was used for sorting using a BD FACS Melody (BD Biosciences) with a 100uM nozzle. Protocol CG000338 from 10x Genomics was used for Chromium Next GEM Single Cell Multiome ATAC + Gene Expression. Briefly, following nuclei isolation, permeabilization was performed, followed by transposition, GEM generation and barcoding using ChipJ (10x Genomics; PN1000234), post-GEM clean up, pre-amplification PCR, cDNA amplification, library construction, and sequencing. Gene expression and ATAC libraires were sequenced to a read depth of 50,000 and 25,000 respectively using a NovaSeq 6000 platform (Illumina) at the McDonnel Genome Institute.

### Multiome data processing

Raw fastq files were aligned to the human GRCh38 reference genome (v) using CellRanger ARC (10x Genomics, v6.1). ArchR^89^ (https://www.archrproject.com) was used to process the ATAC fragments and Seurat was used to process RNA. Quality control was performed to keep nuclei with the following: TSS enrichment > 2, nFrags > 1000, 200 < nUMI GEX < 50,000, and percent mito < 5%. Post-QC nuclei were used for doublet removal in ArchR (ATAC information) and then using scrublet (RNA information). Raw RNA counts were normalized and scaled using SCTransform^90^ regressing out percent mitochondrial reads and nCount_RNA. Principal component analysis, harmony batch integration (by sample), nearest neighbor clustering, and UMAP embedding construction was then performed in Seurat. Cell types were annotated using different expression and knowledge of canonical gene markers. The RNA annotations and normalized gene expression matrix was added to the ArchR project onto the nuclei with the same barcodes. ArchR was used to construct pseudobulk replicates across ATAC clusters and peaks were called using MACS2^91^. The ArchR getMarkerFeatures function was used to identify peaks that are unique to each cell type. The addPeak2GeneLinks function from ArchR was then used to calculate peak to gene links using gene expression and accessibility from the same nucleus with a correlation cut-off of 0.3. To visualize the correspondence between the p2g links, a heatmap was constructed which shows ATAC and RNA z-scores with rows clustered using k-means clustering through the ArchR package plotPeak2GeneHeatmap. We have computed the Pearson correlation coefficient for all genes between the gene expression and accessibility vectors. The ArchR peak 2 gene linkage function was used to identify putative CREs for each gene. In ArchR the addIterativeLSI function was used on the RNA and ATAC modalities respectively with default parameters followed by harmony integration to get modality specific clustering and dimensional reductions. To generate a joint RNA/ATAC embedding, the ArchR addCombinedDims function was used and this combined embedding was used for subsequent UMAP construction and clustering. To compare how the ATAC derived cell clustering compares to the RNA and joint embedding, we generated a confusion matrix.

### LDSC and GWAS Variant Overlap

To test for enrichment of disease risk in the four major cell types, stratified LD score regression was used to partition heritability in cell-type marker peaks^92^. First, cell-type specific marker peaks (FDR < 0.01, log2FC > 1) were obtained using ArchR and converted to GRCh37 coordinates using liftover^93^. GWAS summary statistics for coronary artery disease^8^, diabetes^94^, abdominal aortic aneurysms^95^, diastolic/systolic blood pressure^96^, and stroke^97^ were obtained, and their formats were standardized using MungeSumStats package in R. Stratified LD score analysis was then carried out with --h2-cts flag for cell-type specific analyses. To examine overlap between CAD GWAS variants and ATAC peaks, lead variants from 241 genome-wide significant (p < 5 x 10^-8^) loci and 897 conditionally independent variants meeting FDR cutoff (FDR < 0.01) were obtained from Aragam *et al*^8,98^. Additional variants that are in high LD (r^2^ > 0.80 within 250KB) with these lead variants were obtained from the 1000 Genomes European panel using plink --tag-r2 flag. Additionally, these variants were combined with the functional fine-mapped credible set variants from Aragam et al. to create a superset of GWAS variants [cite Aragam et al.] to be tested for overlap. These variants were overlapped with ATAC peak regions using findOverlap function in IRanges R package.

### Genotyping

To obtain high quality genotypes from patients, DNA from peripheral leukocytes of all individuals was genotyped using Illumina GSA-24-V3 SNP array. Following initial processing using Illumina GenomeStudio software, variants with both 1) minor allele frequency > 5% and call rate < 95% or 2) minor allele frequency < 5% and call rate < 99% were excluded. No individuals were excluded by the call rate exclusion filter (<95%). Additional genotypes were further imputed from the TOPMED panel on the Michigan Imputation Server v. 1.7.1 using minimac4-1.0.2^99^ and phasing with EAGLE^100^. Following imputation, variants with imputation R^2^ < 0.3 and MAF > 0.05 were further filtered for caQTL analysis for a final dataset containing 7,250,405 variants. To conduct combined caQTL mapping, the VCF files from Turner *et al*^34^ were merged with the imputed genotype dataset, and only shared SNPs in two datasets were kept for downstream analysis. Turner *et al*^34^ carried out low coverage whole genome-sequencing; further details are available in the original publication^34^. Following the merge, 5,229,397 variants were available for QTL analysis. Local ancestries were inferred using RMMix2^101^ with default settings. YRI (n=186) and CEU (n=183) from the 1000 Genome Project^102^ were used as AFR / EUR reference populations, respectively.

### Pseudobulk Chromatin Accessibility QTL discovery

Similar to previous studies for single cell caQTL discovery^34,103^, we used RASQUAL^68^ to identify pseudobulk caQTLs in four major cell types (SMC, Endothelial, Myeloid, Fibroblast). RASQUAL maximizes power for caQTL discovery by simultaneously modeling allelic imbalance and total read counts in each locus. To generate fragment counts within each ATAC peak regions, we loaded each fragment object and used Signac’ s FeatureMatrix function to generate a cell-by-fragment count matrix for all cells within each cell type^68^. This matrix was subsequently aggregated at a sample pseudobulk level using rollup function from R slam package. For RASQUAL’s allele-specific modeling, createASVCF.sh script in paired-read mode was used to generate allele-specific fragment counts at each site with additional BAM flag of *-F* 1280 to exclude secondary or PCR duplicate reads. For each cell type, only samples with at least 20 cells of that cell type and only peaks with at least 5 reads on average across all samples were included for the final analysis.

For the caQTL association analysis, all variants within a +/-10KB *cis-*window of the peaks were tested for association. Total library size for each sample was was included as an offset. Age, sex, sequencing site, and first four principal components of genotype data (generated from plink --pca) were included as covariates. For multiple testing correction, we employed a two-step correction procedure recommended by RASQUAL. First, to correct for locus-wide multiple testing, a q-value corresponding to SNP level FDR was calculated using Benjamin-Hochberg method, and the SNP with the lowest q-value was selected for each locus. For multi-locus multiple testing correction, an empirical null distribution was calculated by running RASQUAL four times with --random-permutation flag, which performs association analysis with randomly permuted genotypes. The four runs were averaged to estimate a null-distribution of locus-level q-values, which was subsequently used to obtain q-value cutoffs that would correspond to 1%, 5%, and 10% genome-wide FDR. The caQTL variants were subsequently queried in a database of bulk eQTL datasets using the Qtlizer package^104^.

Pseudobulk caQTLs for the major cell types can be found in Supplementary Tables 14, 16, 18, and 20 at the FDR 5% cut-off. Pseudobulk caQTLs overlapping with GTEx arterial eQTLs at the FDR 5% cut-off can be found in Supplementary Tables 15, 17, 19, and 21.

### Single Cell Chromatin Accessibility QTL Discovery

In addition to the pseudobulk caQTL model, we utilized a Poisson mixed effect (PME) single cell model to map caQTLs in smooth muscle cells. This approach models cell-level fragment counts with random effect covariates to account for relatedness shared by cells from the same sample and batch. Such attempts to model discrete count distributions at a single cell-level has been shown to generally boost power in single-cell QTL discovery compared to pseudobulk and/or linear regression frameworks^48,55,105^. Our PME model was adapted from the Poisson mixed effect model utilized by Nathan *et al*^48^ for single cell eQTL mapping as follows:

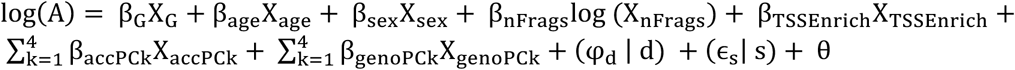

Where ATAC fragment counts (A) was modeled with fixed effect covariates (G=genotype, nFrags=number of fragments per cell, TSSEnrich=TSS enrichment per cell, accPC=principal components of single-cell ATAC counts, genoPC=principal components of genotype data), random effect covariates (d=donor ID, s=sequencing site), and intercept (θ). All quantitative covariates were scaled and centered prior to the regression analysis. Significance of the genotype effect term (β_G_) was tested by likelihood ratio test (LRT) of the full model containing the genotype term and a null model without the genotype term. Due to the high computational load of running Poisson regression analysis on all cells, this analysis was restricted to lead caQTLs variants from the pseudobulk RASQUAL analysis, which models the fragment counts using a negative binomial model. All Poisson models were fit using glmer function from the lme4 R package. We confirmed that our data is generally not over-dispersed by plotting the mean variance relationship between a random subset of peaks. Similarly, utilizing a zero-inflated Poisson regression model or a negative binomial model on a subset of our hits revealed little differences in overall regression estimates. Given that a small proportion of our pseudobulk hits likely represent null results, we did not conduct FDR correction due to our inability to accurately estimate the proportion of results from the null distribution. Instead, we compared Z-values between our pseudobulk and PME models.

A key benefit of modeling individual cells instead of sample-level pseudobulk is our ability to simultaneously model cell-state covariates (e.g. differentiation trajectories, gene/protein expression) as well as potential interaction between genotypes and cell-state covariates. We defined fibromyocyte (FMC) identity score by calculating the gene activity of the top 100 fibromyocyte (modulated SMC) marker genes defined by Wirka *et al*^30^ from our integrated snATAC-seq meta map using ArchR’s addGeneScoreMatrix() function. To test for “dynamic” caQTL where the genotype effect is modified by SMC cell state, we then added the FMC score and a genotype-FMC score interaction term as covariates in the following model:

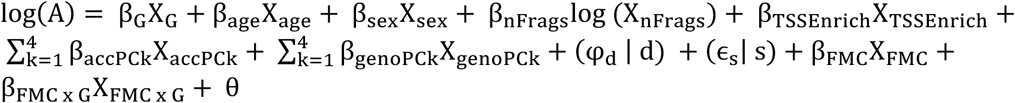

Again, this analysis was carried out just on the significant (FDR<5%) lead variants from the pseudobulk caQTL analyses, though it is entirely possible that there are non-significant pseudobulk caQTLs (i.e. genotype effect is near zero for average FMC score) that may have significant interaction terms. The significance of the interaction term was tested by likelihood ratio test of PME models with and without the interaction term. The LRT p-values were then corrected for multiple testing using the Benjamin-Hochberg method.

Dynamic caQTLs can be found in Supplementary Tables 22.

### scE2G model predictions

We used the scE2G*^Multiome^* model^66^ to predict enhancer–gene connections in 11 cell types from 10x Multiome data (Maya Sheth, Wei-Lin Qiu, Robin Andersson, and Jesse Engreitz, in preparation). In each cell type, scE2G scores every candidate element–gene pair (where candidate elements are ATAC peaks within 5 Mb of the transcription start site of the gene) by integrating several features. Regulatory enhancer–gene interactions were defined as element– gene pairs with a score greater than 0.171.

The scE2G*^Multiome^* model is a supervised classifier adapted for multiomic data from ENCODE-rE2G^46^. It integrates eight features, including 1) a pseudobulk Activity-By-Contact (ABC) score^106^, where 3D contact is estimated by an inverse function of genomic distance; 2) the Kendall correlation across single cells between element accessibility and gene expression; 3) whether the gene is “ubiquitously expressed,” and 4) several other measures of genomic distance and chromatin accessibility around the element and promoter. The score threshold of 0.171 was determined as the score yielding 70% recall when evaluating predictions in K562 cells against CRISPRi-validated enhancer-gene pairs^46^.

scE2G predictions for all cell types can be found in Supplementary Tables 3-13.

### Human coronary artery Hi-C Data Collection and Processing

Bulk *in-situ* Hi-C libraries for 16 different coronary artery samples across a multi-ethnic cohort were generated with the Arima-HiC kit, according to the manufacturer’s protocols. Specifically, one Hi-C library was created from each biological sample, with an input of ∼800k cells each. Restriction enzymes cutting at ^GATC and G^ANTC were utilized in library construction, and approximately 600 million reads were sequenced on an Illumina HiSeq 4000 per library.

To process the data, we used version 2.0.0 of the nf-core^107^ Hi-C pipeline (Zenodo: https://doi.org/10.5281/zenodo.7556794). While we largely used default parameters, we did modify the pipeline by following a recommendation from Arima to trim the first 5 bases from the 5’ end of each adapter sequence, which we accomplished with Cutadapt version 3.4 (https://doi.org/10.14806/ej.17.1.200). Arima recommended this change to help increase the mapping rate when using pipelines such as HiC-Pro^108^, and we did observe an increase in percentage of successfully mapped reads upon implementation. We also set the nf-core pipeline parameter min_mapq to 0, to maximize our retained reads prior to further filtering. Briefly, the pipeline aligned paired-end sequencing reads to human genome hg38, and trimmed unaligned reads with a ligation junction before attempted re-alignment (“rescue” of chimeric reads). Ultimately, ICE-normalized contact maps at 2kb, 5kb, 10kb, 50kb, 100kb, 500kb, and 1Mb resolutions were generated. We assessed maximum library resolution using HiCRes version 2.0^109^, and ultimately decided to focus our analyses on the 10kb maps based on the range of inferred resolutions across samples.

To assess significant loops within each Hi-C library, we used two different algorithms: the cooltools version 0.7.1^110^ “dots” function (based on HiCCUPS^111^), and FitHiC version 2.0.8^112^. For cooltools, we analyzed loops at 2kb, 5kb, and 10kb resolutions, using default parameters for each and donut-based kernels of w=(7, 7, 5) and p=(4, 4, 2), respectively. For FitHiC,we analyzed the same resolutions, and performed a single spline pass per sample, allowing for bias values down to a minimum of 0 based on our observed biases across samples. In both methods, we retained identified loops with q-values ≤ 0.05 as significant. To interrogate topologically associating domains (TADs), we employed cooltools insulation score algorithm, with window sizes of 150kb, 250kb, and 500kb. We kept the boundaries identified as “strong” at each window size to define genomic intervals representing TAD bodies. To visualize genome-wide Hi-C contact maps, we also used cooltools. For visualization of individual loci in concert with gene, loop, and TAD annotations, we used coolbox version 0.3.8^113^. In figures with TAD annotations, TADs across all tested window sizes are shown, whereas significant loop annotations focus on 10kb loop calls only. Intra-chromosomal loop and TAD sizes within each sample were calculated by subtracting the lower bin’s start coordinate from the higher bin’s start coordinate. These data were then visualized using ggplot2.

### HiC network analysis

We kept loops detected in at least 2 patient coronaries for this analysis. To construct an un-biased connected chromatin network^58^, we detected communities of interacting HiC loop anchors using a fast greedy modularity optimization algorithm^59,114^. Using the matrix of connected DNA loops, we used the RBGL package (R interface for Boost graph library algorithms)^115^ – specifically, we used the ftM2graphNEL function to create a graph from the connected matrix and then the connectedComp function to build a connected network graph where each node is a HiC loop and the edges are linking loops to other loops whose anchors overlap. Visualization of the graphs was done using the Rgraphviz package in R. To construct a variant-to-enhancer-to-network (V2E2N) map, we identified loops where any anchor in the network contains a V2E + ENCODE coronary artery H3K27ac peak. Finally, to build a catalogue of all genes predicted to be regulated within a chromatin network, we overlapped all HiC anchors in a network with the E2G to construct a final V2E2N with implicated genes.

### H3K27ac HCASMC and HCAEC HiChIP

HiChIP was adapted from previously published protocols with some modifications^50,116^. Briefly, at least one million human healthy coronary artery smooth muscle cells (HCASMC) (Thermo Scientific, Product code C-017-5C, Lot. 1689414) and human healthy coronary artery endothelial cells (HCAEC) (ATCC, Product code PCS-100-020, Lot 62382179) were crosslinked with 1% Formaldehyde. The initial Hi-C portion followed the Arima HiC protocol described in the Arima High Coverage Hi-C Kit (Arima Genomics, Material Part Number: A410110). For the immunoprecipitation step, sheared chromatin after the Hi-C portion of the protocol was immunoprecipitated with 5 μg of H3K27ac antibody (Abcam, ab4729) with 50 μl of protein G Dynabeads (Invitrogen 10004D) and allowed to incubate overnight at 4°C. The protein G Dynabeads were washed and the immunoprecipitated DNA was de-crosslinked at 67°C for 2 hours and eluted (50 mM sodium bicarbonate, 1% SDS). Subsequently, Dynabeads MyOne Streptavidin C1 beads (Invitrogen 65002) were used to enrich ligated junctions as per Hi-C protocol. Library preparation was performed using the NEB Ultra II library preparation kit (NEB, E7645L), according to the manufacturer’s protocol. 12 PCR cycles were performed using indexed primers and subsequently DNA fragments between 300 to 500 bp were size selected using the Omega MagBind NGS cleanup Magnetic beads (Omega M1378). The libraries were sequenced by paired-end sequencing with at least 300 million read pairs at 2 x 151 bp read length on illumina HiSeq 4000 platforms. Raw paired-end reads in Fastq format for respective samples were processed using the HiCuP pipeline^117^ (Version 0.7.4). The hg38 human genome was used as reference to generate the HiCuP Digest file using two restriction motifs GATC and GANTC via the hicup_digester command for running the HiCuP pipeline. Paired contacts were extracted from aligned filtered bam files using Samtools (Version 1.13) and used as input for the pre function in juicer package to generate .hic files^118^. Paired interaction counts at 5kb resolution were extracted from the matrix, and High-confidence chromatin loops were identified using the FitHiChIP tool as previously described using default parameters^119^. Using HCASMC and HCAEC ChIP-Seq peak files as input, and initial paired interaction files, high-confidence peak-to-all interactions with a 5kb bin size were processed using the bash FitHiChIP_HiCPro.sh script (https://ay-lab.github.io/FitHiChIP/). High-confidence chromatin interactions were sorted and indexed in bed format and converted to long-range format for visualization on the WashU browser. The overlay of HiChIP, ChIP-seq, and GWAS data was plotted using the locuszoomr package (version 0.3.5) and R (version 4.3.1). Raw Fastq files and processed HiChIP data will be uploaded to NCBI SRA upon publication of the manuscript.

### H3K27ac Chromatin Immunoprecipitation (ChIP-Seq)

Chromatin immunoprecipitation with sequencing (ChIP-seq) was performed as previously described with minor optimisations^120^. Briefly, one million human coronary artery smooth muscle cells (HCASMC) and human coronary endothelial cells (HCAEC) from the same lot used for HiChIP experiments were crosslinked with 1% Formaldehyde for 10 min at room temperature and quenched with glycine (125mM) for 5 min. The cells were rinsed twice with cold 1XPBS and sonicated in nuclei lysis buffer (50mM HEPES-KOH, pH 7.5, 150mM NaCl, 1mM EDTA, 1% Triton X, 0.1% Sodium deoxycholate, 1% SDS, Takara, 1x protease inhibitor) using the Bioruptor sonicator to obtain chromatin fragments between 200 to 500 bp. The fragmented chromatin was immunoprecipitated overnight at 4°C, with 5 μg of H3K27ac antibody (Abcam, ab4729) and 50 μl of protein G beads (Invitrogen, 10004D). Subsequently, the beads were washed and immunoprecipitated DNA was de-crosslinked and eluted in elution buffer (50 mM Tris-HCI, pH7.5, and 10 mM EDTA). Eluted ChIP fragments were isolated using Phenol-Chloroform extraction method, and DNA was purified by ethanol precipitation. Library preparation was performed on the eluted ChIP DNA using the NEB Ultra II library preparation kit, according to the manufacturer’s protocol. 10 PCR cycles were performed using indexed primers and the library of DNA fragments with sizes between 300 to 500 bp was selected using the Omega MagBind NGS cleanup Magnetic beads (Omega M1378). The libraries were sent for paired end sequencing with at least 50 million read pairs at 2 x 151 bp read length on the illumina HiSeq 4000 platform. Paired-end sequencing reads were aligned to human genome (hg38) using BWA mem version 0.7.5^121^, and reads were deduplicated using Picard MarkDuplicates function. Aligned mapped reads in BAM file were used as input for peak-calling using Dfilter^122^ to identify significant H3K27ac peaks using the following setting: -ks=60, -bs=100, -Ipvalue=8. Bigwig tracks were normalized for sequencing depths and the sum of normalized binned tag-count represents the peak height. Raw Fastq files and processed ChIP-seq data will be uploaded to NCBI SRA upon manuscript publication.

### Culture of HCAEC and HCASMC primary cells

human healthy coronary artery smooth muscle cells (HCASMC) (Thermo Scientific, Product code C-017-5C, Lot. 1689414) are cultured in human vascular smooth muscle (VSMC) basal medium (Gibco, M231500), in Smooth Muscle Growth Supplement (SMGS) (Gibco, S00725) and 1% penicillin/streptomycin. human healthy coronary artery endothelial cells (HCAEC) (ATCC, Product code PCS-100-020, Lot 62382179) are cultured in Endothelial Cell Growth medium MV2 (PromoCell, C-22022).

### Construction of lentiviral sgRNA plasmids and Lentivirus production

The Lentiguide-Puro plasmid was obtained as a gift from Feng Zhang (Addgene #52963, LentiGuide-Puro) in which the Non-Targeting Control sgRNAs and sgRNAs targeting the TSS and Regulatory enhancer were annealed and ligated into the Esp3I cut-site of the Lentiguide-Puro vector using T4 Ligase (NEB, M0202) as per manufacturer’s instructions. Ligation products were transformed into One Shot Stbl3 Chemically competent E.coli (Invitrogen, C737303), and plasmids of positive clones were isolated using FavorPrep Plasmid DNA Extract Kit (Favorgen, FAPDE001-1). The CRISPR interference expression vector for Lenti-dCas9-KRAB-blast (Addgene, #89567) was a gift from Gary Hon. For lentivirus production, HEK293T cells were cultured in DMEM+10% FBS until 70% confluency prior to transfection. 10ug of individual plasmids, 7.5ug of pMDLg/RRE, 2.5ug of pRSV-Rev and 2.5ug of pMD2.G lentivirus packaging plasmids were co-transfected with 50ul of Polyethylenimine (PEI) diluted in 3ml of Opti-MEM™ I Reduced Serum Medium (#31985070, ThermoFisher Scientific). Medium was refreshed overnight with reduced 5% FBS in DMEM medium, and the expended medium was collected twice after 24hr and 48 hr respectively. Pooled supernatant was filtered through a PES 0.45uM filter and viral particles were concentrated using Viro-PEG Lentivirus Concentrator (Ozbiosciences, #LVG100) as per manufacturer’s instructions.

The sgRNAs are listed below:

Non-Targeting Control_sgRNA_F: CACCGAACGTGCTGACGATGCGGGC
Non-Targeting Control_sgRNA_R: AAACGCCCGCATCGTCAGCACGTTC
AMOTL2_TSS_sgRNA_F: 5’-CACCGGCGCGAACAGCCAGAGCGT-3’
AMOTL2_TSS_sgRNA_R: 5’-AAACACGCTCTGGCTGTTCGCGCC-3’
AMOTL2_Enhancer_sgRNA1_F: 5’-CACCGTATTCATAGACATCACTAA-3’
AMOTL2_Enhancer_sgRNA1_R: 5’-AAACTTAGTGATGTCTATGAATAC-3’
AMOTL2_Enhancer_sgRNA2_F: 5’-CACCGATCCCTATGGAATCCTTGG-3’
AMOTL2_Enhancer_sgRNA2_R: 5’-AAACCCAAGGATTCCATAGGGATC-3’

### Transduction of primary cells for CRISPR interference and gene expression analysis

At least two independent sgRNAs targeting a single regulatory element were tested against the non-targeting control. sgRNA targeting the transcription start site (TSS) was used as positive control. Both human healthy coronary artery smooth muscle cells (HCASMC) (Thermo Scientific, Product code C-017-5C, Lot. 1689414) and human healthy coronary artery endothelial cells (HCAEC) (ATCC, Product code PCS-100-020, Lot 62382179) were co-transduced in 5ug/ml polybrene with lentiviral particles packaged from Lenti-dCas9-KRAB-blast (Addgene, #89567)^123^, and individual sgRNAs cloned into Lentiguide-Puro backbone (Addgene, #52963)^124^ at multiplicity of infection of 3. Respective HCASMC and HCAEC growth media were refreshed after 24h of infection, and co-selection with 10ug/ml blasticidin (Gibco, A1113903), and 1ug/ml puromycin (Gibco, A1113803) was performed after 72h post-infection. Selected cells were allowed to recover and expanded for 2 weeks before RNA isolation using Trizol Reagent (Gibco, 15596026) and Direct-Zol RNA miniprep (Zymo Research, R2052) as per manufacturer’s instructions. cDNA conversion was performed with at least 200ng RNA using HiScript III RT SuperMix (Vazyme Biotech, R323-01) and RT-PCR was performed using gene-specific primers to assess for gene expression.

Below are the following RT-PCR primers used:

AMOTL2_F: AGTGAGCGACAAACAGCAGACG
AMOTL2_R: ATCTCTGCTCCCGTGTTTGGCA
ANAPC13_F: GATTGATGATGCTTGGCG
ANAPC13_R: GTAAGGCTAAGTCTGTCC
CEP63_F: TGGGAAGGACGTACACATGC
CEP63_R: ACATCCAACTGACTCCTAAGACT
GAPDH_F: GTGGACCTGACCTGCCGTCT
GAPDH_R: GGAGGAGTGGGTGTCGCTGT
PPIA_F: CACCGTGTTCTTCGACATTG
PPIA_R: TTCTGCTGTCTTTGGGACCT

## Data Availability

Raw and processed sequencing files can be found on the Gene Expression Omnibus super series ().

## Code Availability

Scripts used for analysis in this manuscript can be found at (https://github.com/jamrute).

---

**Extended Data Figure 1.**
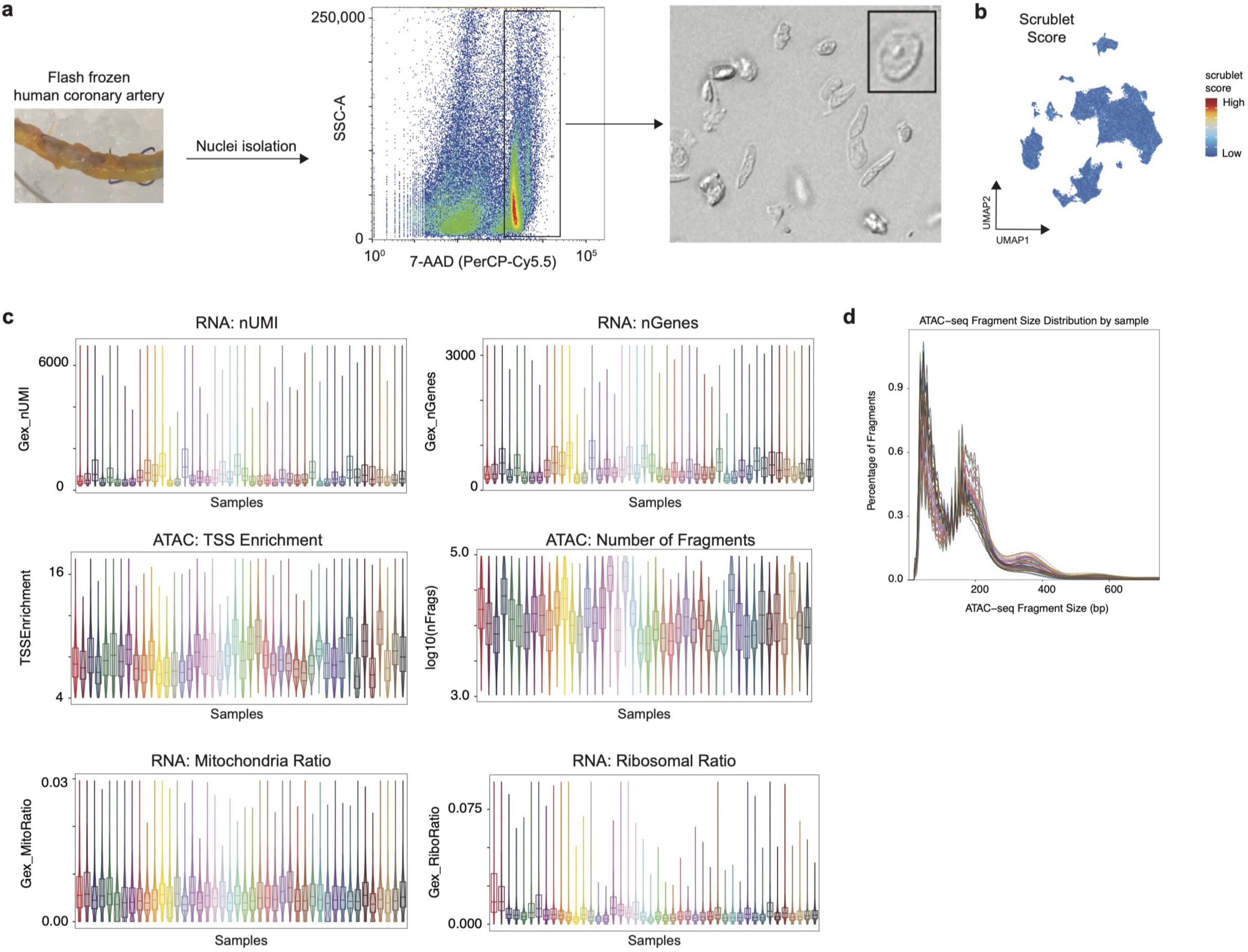
Multiome quality control. (a) Experimental workflow to isolate nuclei from flash frozen coronary arteries with flow cytometry to sort for 7-AAD+ nuclei with final morphology. (b) Scrublet score after doublet removal. (c) Quality control metrics for RNA (number of UMI counts, number of genes, mitochondrial RNA ratio, and ribosomal RNA ratio per nucleus) and ATAC (TSS enrichment and number of fragments per nucleus) grouped by sample. (d) snATAC-seq fragment size distribution per sample.

**Extended Data Figure 2.**
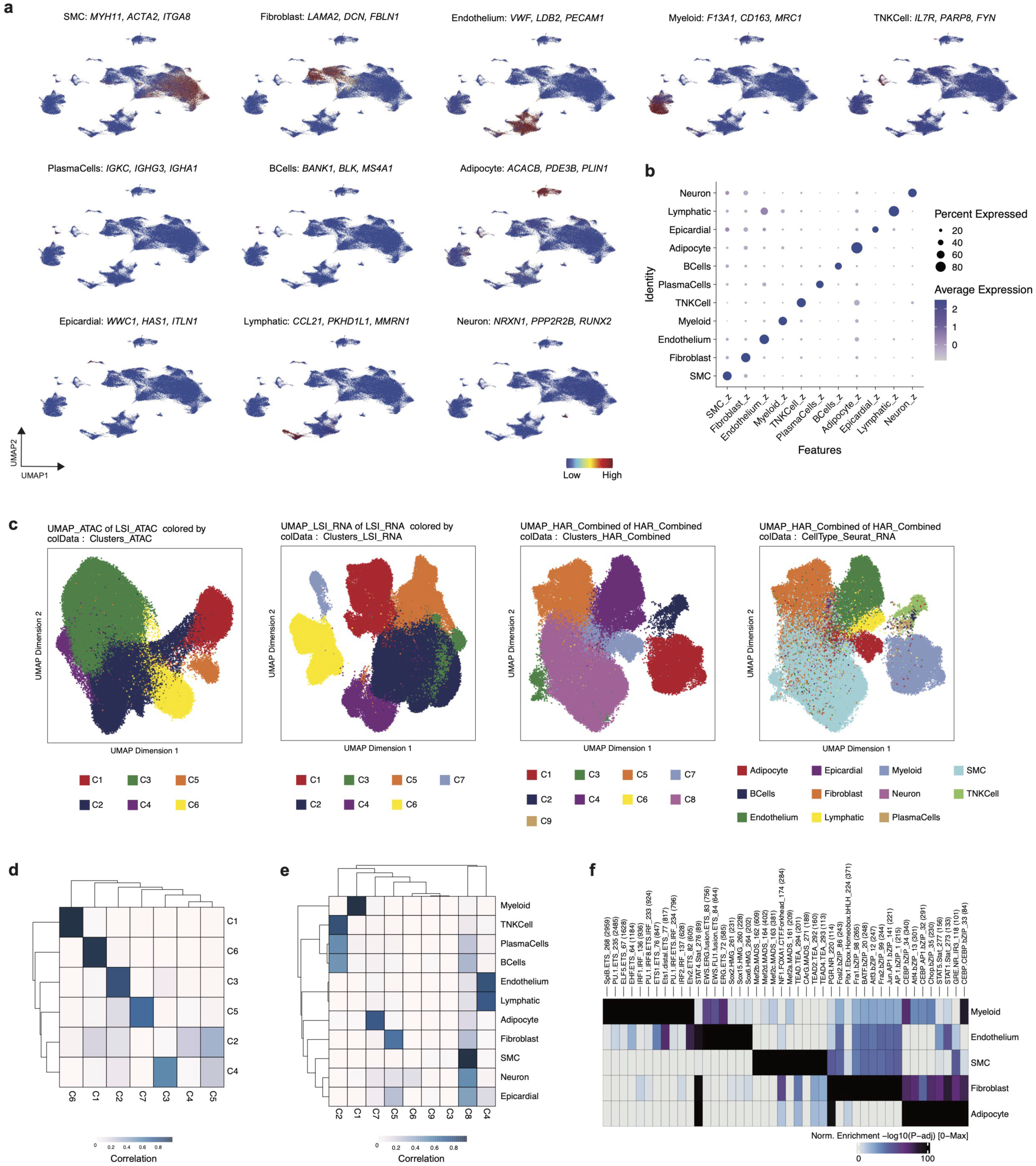
Multimodal clustering of Multiome data. (a) Gene set scores for marker genes (adjusted p-value < 0.05 and log_2_FC > 0.58) in each cell type. (b) DotPlot of marker gene set score form (a) grouped by cell type. (c) UMAP embedding and clustering using ATAC LSI, RNA LSI, RNA/ATAC integrated with harmony batch correction, and RNA/ATAC integrated with harmony batch correction with annotations from RNA based clustering. Confusion matrix for cluster annotations in (d) RNA and ATAC clustering and (e) RNA/ATAC integrated with harmony batch correction and RNA. (f) Motif enrichment using chromVar by major cell types using differentially accessible peaks (FDR < 0.1 and log_2_FC > 0.5).

**Extended Data Figure 3.**
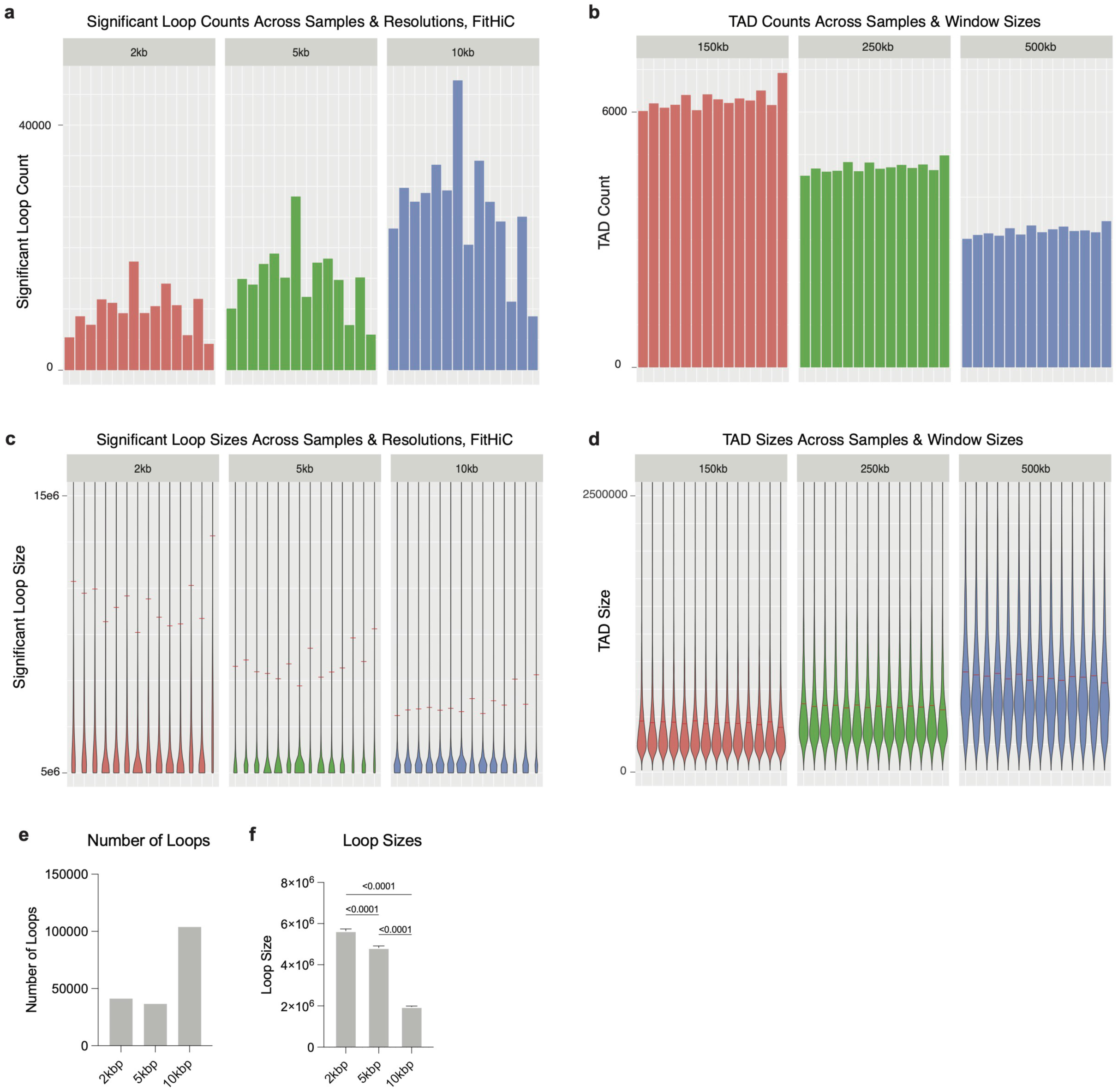
HiC quality control. Number of significant (a) loops called b FitHiC and (b) TADs identified at different resolutions by sample. (c) Loop and (d) TAD size distributions by sample. (e) Number of loops called using FitHiC at various anchor resolutions. (f) Average loop size at various anchor resolutions.

**Extended Data Figure 4.**
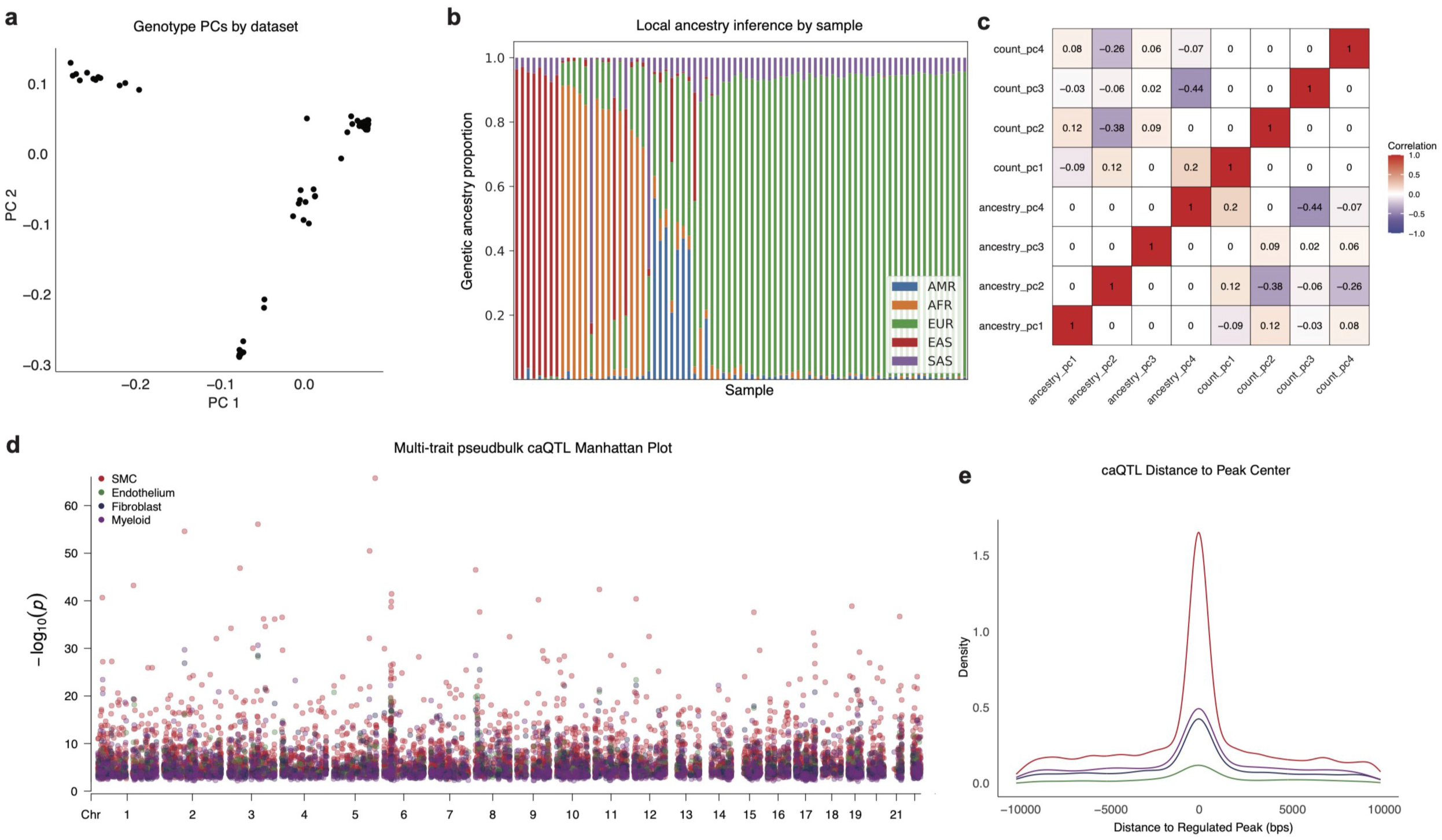
Genotyping and pseudobulk caQTL discovery. (a) PC1 and 2 for genotyping data (post imputation from the TOPMED panel and filtering for variants with imputation R^2^ < 0.3 and MAF > 0.05) panel for each sample. (b) Local ancestry inference (YRI (n=186) and CEU (n=183) from the 1000 Genome Project^102^ were used as AFR / EUR reference populations, respectively.) genetic proportion stack plot by sample. (c) Correlation heatmap for ancestry PCs and ATAC count PCs. (d) Multi-trait pseudobulk Manhattan plot for caQTL colored by cell type genome wide. (e) Distance from caQTL to the regulated peak colored by cell type.

**Extended Data Figure 5.**
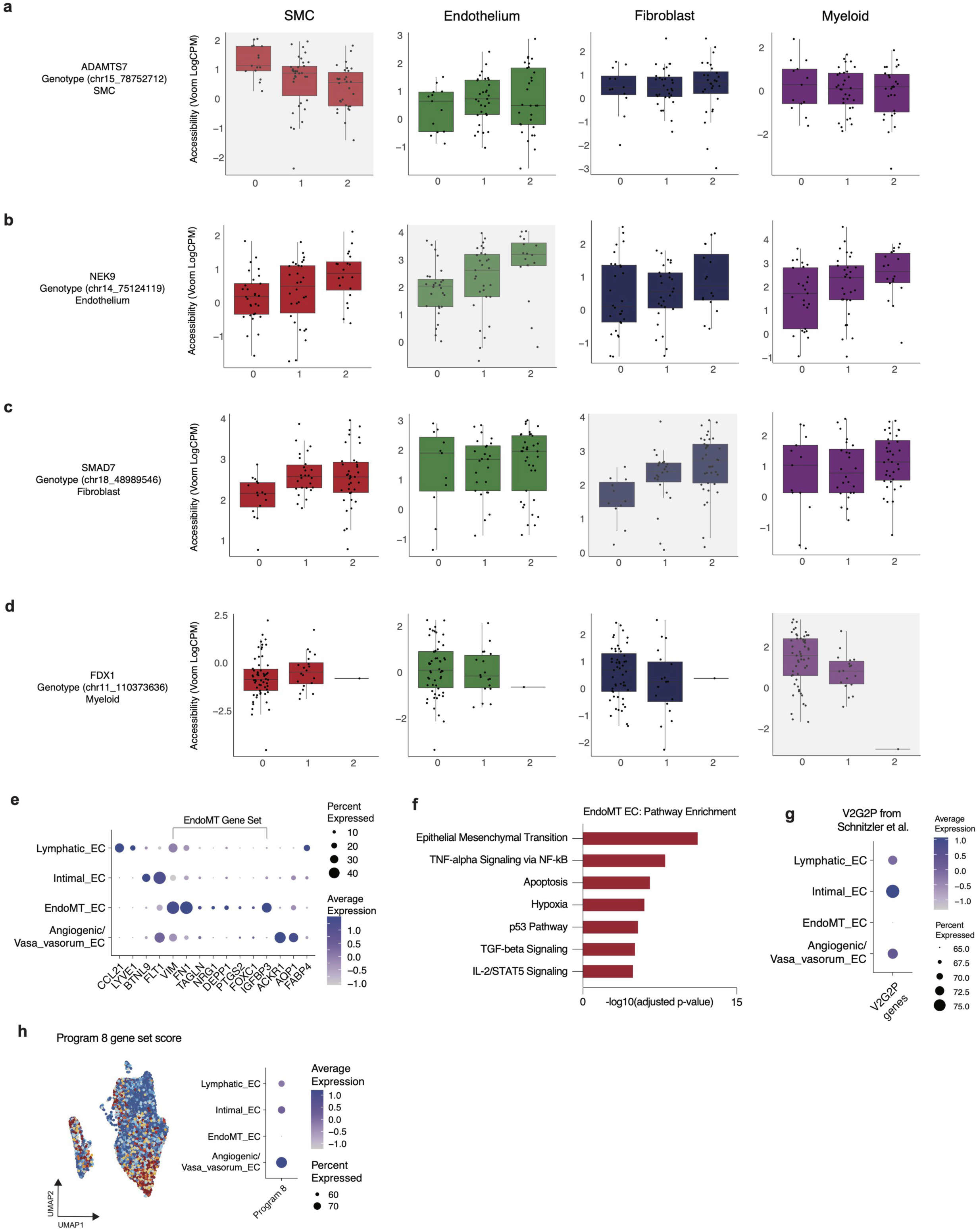
Cell type and state regulation of molecular traits. Pseudobulk accessibility by genotype plots for (a) *ADAMTS7*, (b) *NEK9*, (c) *SMAD7*, and (d) *FDX* are cell type specific caQTLs for SMCs, endothelium, fibroblasts and myeloid cells respectively. (e) DotPlot of canonical marker genes for endothelium cell biology grouped by mapped cell state annotation. (f) Pathway enrichment for statistically significant genes upregulated in EndoMT (adjusted p-value < 0.05 and log2FC > 0.58). (g) Endothelium acting loci from V2G2P in Schnitzler *et al*^47^ gene set score grouped by EC state. (h) Program 8 gene set score from Schnitzler *et al*^47^ in UMAP embedding plot (left) and DotPlot grouped by EC state (right).

**Extended Data Figure 6.**
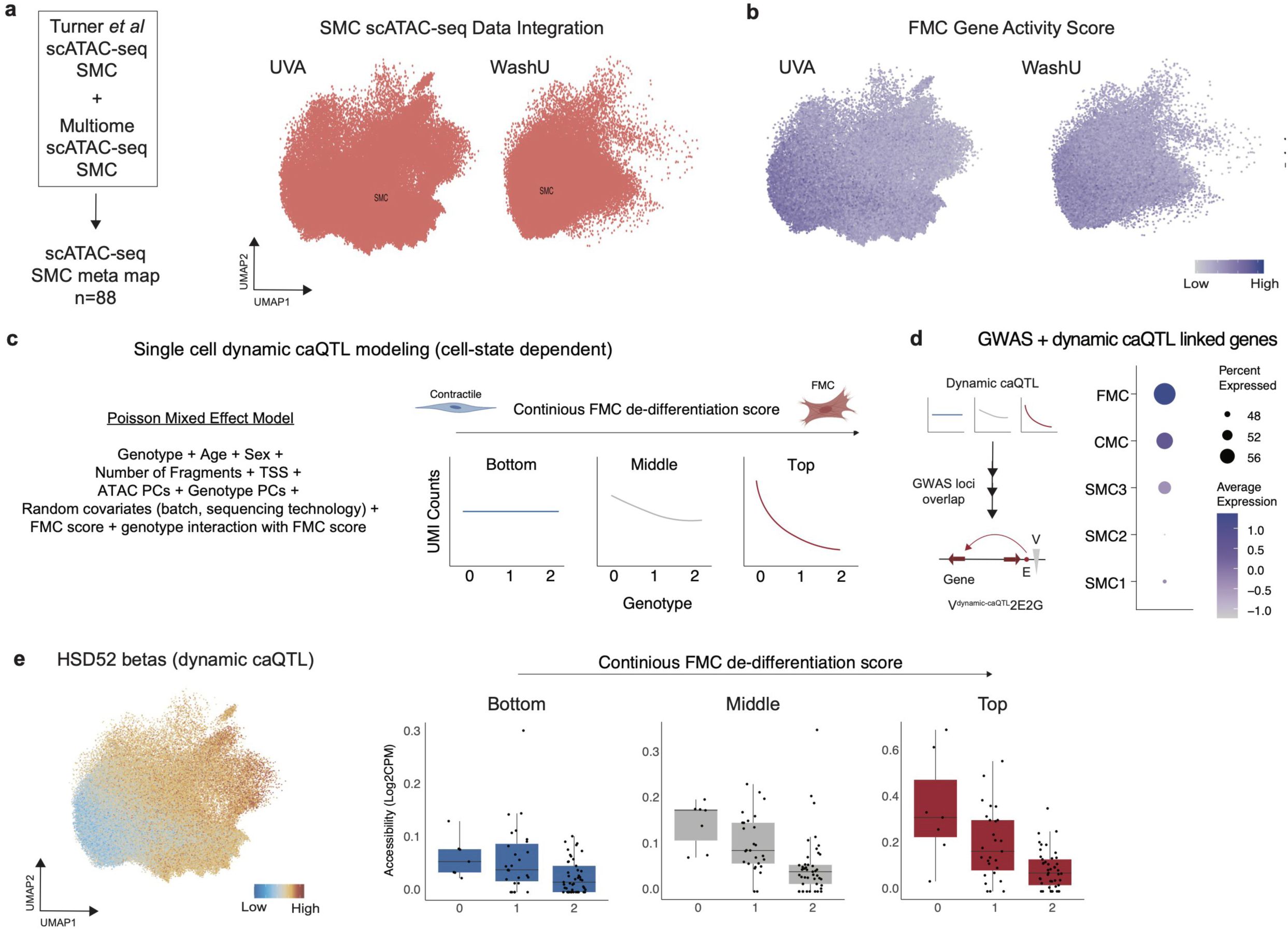
Dynamic caQTL modeling. (a) SMCs integrated from Turner *et al*^34^ and Multiome snATAC-seq to construct a meta-map for dynamic single cell QTL modeling grouped by dataset. (b) FMC marker genes gene activity score split by dataset. (c) Framework for single cell dynamic caQTL modeling with a Poisson Mixed Effect model showing an FMC state dependent effect of genotype on chromatin accessibility. (d) Gene set score for dynamic caQTLs overlapped with CAD GWAS variants linked to genes with scE2G grouped by SMC state. (e) *HSD52* betas for dynamic caQTL at rs658956 in UMAP embedding (left) and pseudobulk box plots by genotype and FMC score tertile (right).

**Extended Data Figure 7.**
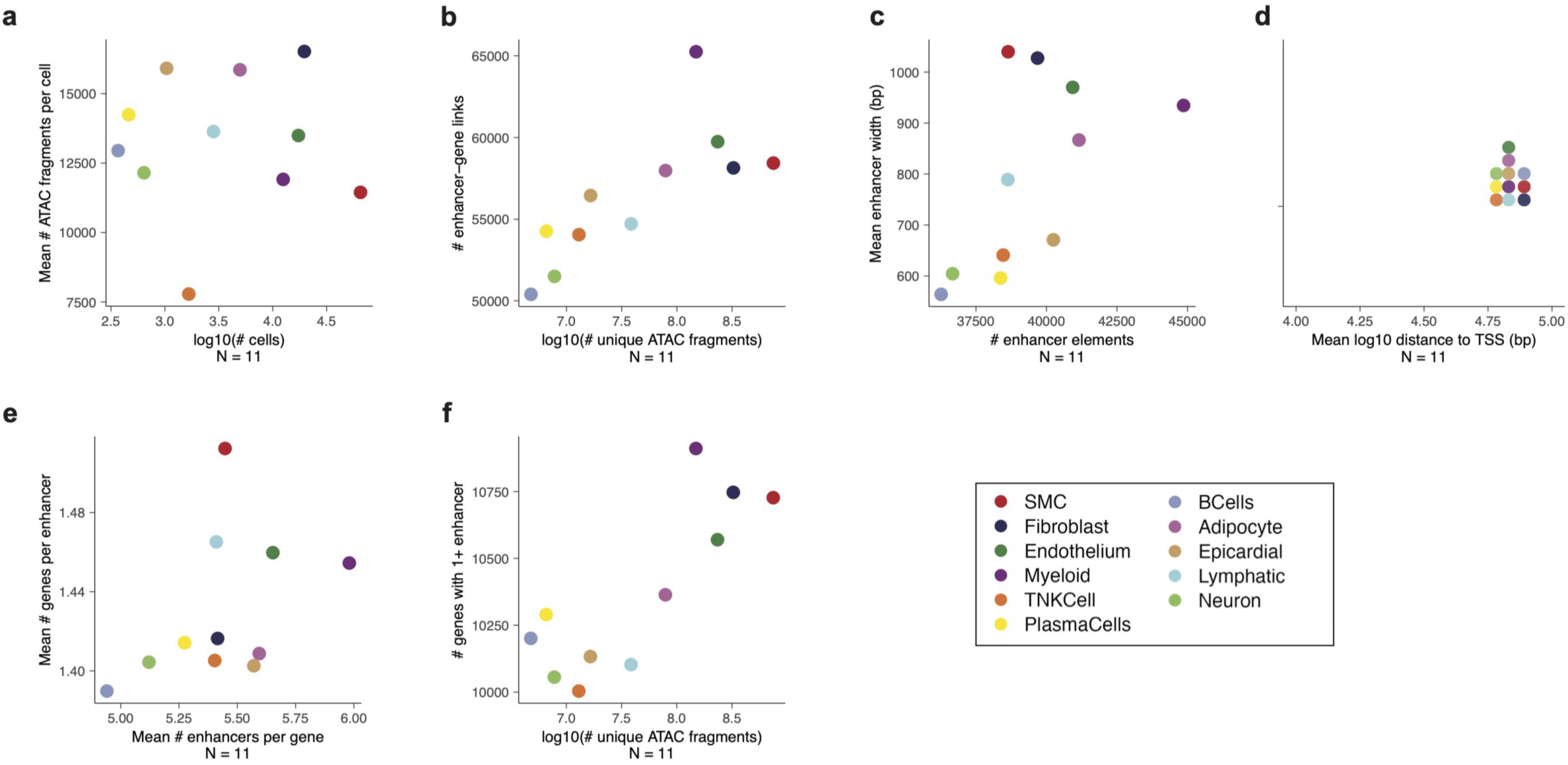
Each point represents enhancer-gene predictions by scE2G*^Multiome^* in one cell cluster (N = 11). All properties were computed on binarized predictions omitting promoter elements. (a) Average number of unique ATAC fragments per cell by number of cells per cluster. (b) Number of enhancer-gene links by number of unique ATAC fragments per cluster. T (c) Mean enhancer width by number of unique enhancer elements regulating at least one distal gene. (d) Mean distance between enhancer element and target gene transcription start site. (e) Mean number of enhancers per gene by mean number of genes per enhancer. (f) Number of genes with at least one non-promoter enhancer by number of unique ATAC fragments.

## References

1. Khera, A. V. et al. Genome-wide polygenic scores for common diseases identify individuals with risk equivalent to monogenic mutations. Nature Genetics 2018 50:9 50, 1219–1224 (2018).

2. Uffelmann, E. et al. Genome-wide association studies. Nature Reviews Methods Primers 2021 1:1 1, 1–21 (2021).

3. Martin, S. S. et al. 2024 Heart Disease and Stroke Statistics: A Report of US and Global Data From the American Heart Association. Circulation 149, E347–E913 (2024).

4. Kathiresan, S. & Srivastava, D. Leading Edge Review Genetics of Human Cardiovascular Disease. (2012) doi:10.1016/j.cell.2012.03.001.

5. Erdmann, J., Kessler, T., Munoz Venegas, L. & Schunkert, H. A decade of genome-wide association studies for coronary artery disease: the challenges ahead. Cardiovasc Res 114, 1241–1257 (2018).

6. Khera, A. V. & Kathiresan, S. Genetics of coronary artery disease: discovery, biology and clinical translation. Nat Rev Genet 18, 331–344 (2017).

7. Musunuru, K. & Kathiresan, S. Genetics of Common, Complex Coronary Artery Disease. Cell 177, 132–145 (2019).

8. Aragam, K. G. et al. Discovery and systematic characterization of risk variants and genes for coronary artery disease in over a million participants. Nature Genetics 2022 54:12 54, 1803–1815 (2022).

9. Tcheandjieu, C. et al. Large-scale genome-wide association study of coronary artery disease in genetically diverse populations. Nature Medicine 2022 28:8 28, 1679–1692 (2022).

10. Shapiro, M. D., Tavori, H. & Fazio, S. PCSK9: From Basic Science Discoveries to Clinical Trials. Circ Res 122, 1420 (2018).

11. Abifadel, M. et al. Mutations in PCSK9 cause autosomal dominant hypercholesterolemia. Nature Genetics 2003 34:2 34, 154–156 (2003).

12. Ridker, P. M. et al. Inflammation, Cholesterol, Lipoprotein(a), and 30-Year Cardiovascular Outcomes in Women. New England Journal of Medicine (2024) doi:10.1056/NEJMOA2405182/SUPPL_FILE/NEJMOA2405182_DATA-SHARING.PDF.

13. Karalis, D. G. Intensive Lowering of Low-Density Lipoprotein Cholesterol Levels for Primary Prevention of Coronary Artery Disease. Mayo Clin Proc 84, 345 (2009).

14. Bayturan, O. et al. Clinical Predictors of Plaque Progression Despite Very Low Levels of Low-Density Lipoprotein Cholesterol. J Am Coll Cardiol 55, 2736–2742 (2010).

15. Hartmann, K., Seweryn, M. & Sadee, W. Interpreting coronary artery disease GWAS results: A functional genomics approach assessing biological significance. PLoS One 17, (2022).

16. Kessler, T. & Schunkert, H. Coronary Artery Disease Genetics Enlightened by Genome-Wide Association Studies. JACC Basic Transl Sci 6, 610–623 (2021).

17. Kamb, A., Harper, S. & Stefansson, K. Human genetics as a foundation for innovative drug development. Nat Biotechnol 31, 975–978 (2013).

18. Dowden, H. & Munro, J. Trends in clinical success rates and therapeutic focus. Nat Rev Drug Discov 18, 495–496 (2019).

19. Haldar, S. M. Keeping translational research grounded in human biology. J Clin Invest 134, (2024).

20. Wang, X., Savic, D., Hovig, E., Trynka, G. & Cano-Gamez, E. Making sense of GWAS: using epigenomics and genome engineering to understand the functional relevance of SNPs in non-coding regions of the human genome. Epigenetics & Chromatin 2015 8:1 8, 1–18 (2015).

21. Cano-Gamez, E. & Trynka, G. From GWAS to Function: Using Functional Genomics to Identify the Mechanisms Underlying Complex Diseases. Front Genet 11, 505357 (2020).

22. Mardis, E. R. The impact of next-generation sequencing technology on genetics. Trends Genet. 24, 133–141 (2008).

23. Schuster, S. C. Next-generation sequencing transforms today’s biology. Nat. Methods 5, 16–18 (2008).

24. Tang, F. et al. mRNA-Seq whole-transcriptome analysis of a single cell. Nature Methods 2009 6:5 6, 377–382 (2009).

25. Kolodziejczyk, A. A., Kim, J. K., Svensson, V., Marioni, J. C. & Teichmann, S. A. The Technology and Biology of Single-Cell RNA Sequencing. Mol Cell 58, 610–620 (2015).

26. Koenig, A. L. et al. Single-cell transcriptomics reveals cell-type-specific diversification in human heart failure. Nature Cardiovascular Research 2022 1:3 1, 263–280 (2022).

27. Kuppe, C. et al. Spatial multi-omic map of human myocardial infarction. Nature (2022) doi:10.1038/s41586-022-05060-x.

28. Tucker, N. R. et al. Transcriptional and Cellular Diversity of the Human Heart. Circulation 142, 466–482 (2020).

29. Chaffin, M. et al. Single-nucleus profiling of human dilated and hypertrophic cardiomyopathy. Nature (2022) doi:10.1038/S41586-022-04817-8.

30. RC, W., et al. Atheroprotective roles of smooth muscle cell phenotypic modulation and the TCF21 disease gene as revealed by single-cell analysis. Nat Med 25, 1280–1289 (2019).

31. Zhao, Q. et al. A cell and transcriptome atlas of the human arterial vasculature. bioRxiv 2024.09.10.612293 (2024) doi:10.1101/2024.09.10.612293.

32. Amrute, J. M. et al. Targeting immune–fibroblast cell communication in heart failure. Nature 2024 1–11 (2024) doi:10.1038/s41586-024-08008-5.

33. Amrute, J. M. et al. Defining cardiac functional recovery in end-stage heart failure at single-cell resolution. Nature cardiovascular research 2, 399–416 (2023).

34. Turner, A. W. et al. Single-nucleus chromatin accessibility profiling highlights regulatory mechanisms of coronary artery disease risk. Nature Genetics 2022 54:6 54, 804–816 (2022).

35. Smith, E. L., Mok, G. F. & Münsterberg, A. Investigating chromatin accessibility during development and differentiation by ATAC-sequencing to guide the identification of cis-regulatory elements. Biochem Soc Trans 50, 1167 (2022).

36. Sun, Y., Miao, N. & Sun, T. Detect accessible chromatin using ATAC-sequencing, from principle to applications. Hereditas 156, 29 (2019).

37. Klemm, S. L., Shipony, Z. & Greenleaf, W. J. Chromatin accessibility and the regulatory epigenome. Nat Rev Genet 20, 207–220 (2019).

38. Duren, Z. et al. Regulatory analysis of single cell multiome gene expression and chromatin accessibility data with scREG. Genome Biol 23, 1–19 (2022).

39. Wang, S. K. et al. Single-cell multiome of the human retina and deep learning nominate causal variants in complex eye diseases. Cell Genomics 2, 100164 (2022).

40. Mitra, S. et al. Single-cell multi-ome regression models identify functional and disease-associated enhancers and enable chromatin potential analysis. Nature Genetics 2024 56:4 56, 627–636 (2024).

41. Zhu, K. et al. Multi-omic profiling of the developing human cerebral cortex at the single-cell level. Sci Adv 9, (2023).

42. Badia-i-Mompel, P. et al. Gene regulatory network inference in the era of single-cell multi-omics. Nat Rev Genet 24, 739–754 (2023).

43. Mathys, H. et al. Single-cell multiregion dissection of Alzheimer’s disease. Nature 2024 632:8026 632, 858–868 (2024).

44. Xiong, X. et al. Epigenomic dissection of Alzheimer’s disease pinpoints causal variants and reveals epigenome erosion. Cell 186, 4422–4437.e21 (2023).

45. Ma, S. et al. Chromatin Potential Identified by Shared Single-Cell Profiling of RNA and Chromatin. Cell 183, 1103–1116.e20 (2020).

46. Gschwind, A. R., et al. An encyclopedia of enhancer-gene regulatory interactions in the human genome. bioRxiv (2023) doi:10.1101/2023.11.09.563812.

47. Schnitzler, G. R. et al. Convergence of coronary artery disease genes onto endothelial cell programs. Nature 626, 799–807 (2024).

48. Nathan, A., et al. Single-cell eQTL models reveal dynamic T cell state dependence of disease loci. | Nature | 606, (2022).

49. Rao, S. S. P. et al. A three-dimensional map of the human genome at kilobase resolution reveals principles of chromatin looping. Cell 159, 1665 (2014).

50. Mumbach, M. R. et al. HiChIP: efficient and sensitive analysis of protein-directed genome architecture. Nat Methods 13, 919–922 (2016).

51. Gate, R. E. et al. Genetic determinants of co-accessible chromatin regions in activated T cells across humans. Nature Genetics 2018 50:8 50, 1140–1150 (2018).

52. Degner, J. F. et al. DNase I sensitivity QTLs are a major determinant of human expression variation. Nature 2012 482:7385 482, 390–394 (2012).

53. Xiong, X. et al. Epigenomic dissection of Alzheimer’s disease pinpoints causal variants and reveals epigenome erosion. Cell 186, 4422 (2023).

54. Cuomo, A. S. E., Nathan, A., Raychaudhuri, S., MacArthur, D. G. & Powell, J. E. Single-cell genomics meets human genetics. Nat Rev Genet 24, 535 (2023).

55. Fitzgerald, T., Jones, A. & Engelhardt, B. E. A Poisson reduced-rank regression model for association mapping in sequencing data. BMC Bioinformatics 23, (2022).

56. Luo, Y. et al. New developments on the Encyclopedia of DNA Elements (ENCODE) data portal. Nucleic Acids Res 48, D882–D889 (2020).

57. Dunham, I. et al. An integrated encyclopedia of DNA elements in the human genome. Nature 489, 57–74 (2012).

58. Phanstiel, D. H. et al. Static and Dynamic DNA Loops form AP-1-Bound Activation Hubs during Macrophage Development. Mol Cell 67, 1037–1048.e6 (2017).

59. Rustamaji, H. C., Kusuma, W. A., Nurdiati, S. & Batubara, I. Community detection with Greedy Modularity disassembly strategy. Scientific Reports 2024 14:1 14, 1–17 (2024).

60. Nikpay, M. et al. A comprehensive 1000 Genomes–based genome-wide association meta-analysis of coronary artery disease. Nature Genetics 2015 47:10 47, 1121–1130 (2015).

61. Elmentaite, R., Domínguez Conde, C., Yang, L. & Teichmann, S. A. Single-cell atlases: shared and tissue-specific cell types across human organs. Nat. Rev. Genet. 23, 395–410 (2022).

62. Litviňuková, M. et al. Cells of the adult human heart. 588, 466–472 (2020).

63. Reichart, D. et al. Pathogenic variants damage cell composition and single-cell transcription in cardiomyopathies. Science (1979) 377, (2022).

64. Kartha, V. K. et al. Functional inference of gene regulation using single-cell multi-omics. Cell Genomics 2, 100166 (2022).

65. Baysoy, A., Bai, Z., Satija, R. & Fan, R. The technological landscape and applications of single-cell multi-omics. Nature Reviews Molecular Cell Biology 2023 24:10 24, 695–713 (2023).

66. GitHub - EngreitzLab/sc-E2G: Pipeline to run sc-E2G. https://github.com/EngreitzLab/sc-E2G.

67. Aguet, F. et al. The GTEx Consortium atlas of genetic regulatory effects across human tissues. Science (1979) 369, 1318–1330 (2020).

68. Kumasaka, N., Knights, A. J. & Gaffney, D. J. Fine-mapping cellular QTLs with RASQUAL and ATAC-seq. Nat Genet 48, 206–213 (2016).

69. Alencar, G. F. et al. Stem Cell Pluripotency Genes Klf4 and Oct4 Regulate Complex SMC Phenotypic Changes Critical in Late-Stage Atherosclerotic Lesion Pathogenesis. Circulation 2045–2059 (2020) doi:10.1161/CIRCULATIONAHA.120.046672.

70. Allahverdian, S., Chaabane, C., Boukais, K., Francis, G. A. & Bochaton-Piallat, M. L. Smooth muscle cell fate and plasticity in atherosclerosis. Cardiovasc Res 114, 540 (2018).

71. Zhao, Y. et al. “Stripe” transcription factors provide accessibility to co-binding partners in mammalian genomes. Mol Cell 82, 3398–3411.e11 (2022).

72. Gehrke, A. R. et al. Acoel genome reveals the regulatory landscape of whole-body regeneration. Science (1979) 363, (2019).

73. Schleif, R. DNA looping. Annu Rev Biochem 61, 199–223 (1992).

74. Matthews, K. S. DNA looping. Microbiol Rev 56, 123 (1992).

75. Hansen, A. S., Cattoglio, C., Darzacq, X. & Tjian, R. Recent evidence that TADs and chromatin loops are dynamic structures. Nucleus 9, 20 (2018).

76. Grubert, F. et al. Landscape of cohesin-mediated chromatin loops in the human genome. Nature 2020 583:7818 583, 737–743 (2020).

77. Xu, J. et al. Subtype-specific 3D genome alteration in acute myeloid leukaemia. Nature 2022 611:7935 611, 387–398 (2022).

78. Heffel, M. G. et al. Temporally distinct 3D multi-omic dynamics in the developing human brain. Nature 2024 17, 1–9 (2024).

79. Lambuta, R. A. et al. Whole-genome doubling drives oncogenic loss of chromatin segregation. Nature 2023 615:7954 615, 925–933 (2023).

80. Nasser, J. et al. Genome-wide enhancer maps link risk variants to disease genes. Nature 2021 593:7858 593, 238–243 (2021).

81. Flavahan, W. A. et al. Altered chromosomal topology drives oncogenic programs in SDH-deficient GISTs. Nature 2019 575:7781 575, 229–233 (2019).

82. Erdmann-Pham, D. D. et al. Tracing cancer evolution and heterogeneity using Hi-C. Nature Communications 2023 14:1 14, 1–17 (2023).

83. Yang, J. et al. Analysis of chromatin organization and gene expression in T cells identifies functional genes for rheumatoid arthritis. Nature Communications 2020 11:1 11, 1–13 (2020).

84. Ron, G., Globerson, Y., Moran, D. & Kaplan, T. Promoter-enhancer interactions identified from Hi-C data using probabilistic models and hierarchical topological domains. Nature Communications 2017 8:1 8, 1–12 (2017).

85. Schöpflin, R. et al. Integration of Hi-C with short and long-read genome sequencing reveals the structure of germline rearranged genomes. Nature Communications 2022 13:1 13, 1–15 (2022).

86. Ing-Simmons, E. et al. Independence of chromatin conformation and gene regulation during Drosophila dorsoventral patterning. Nature Genetics 2021 53:4 53, 487–499 (2021).

87. Schmitt, A. D. et al. A Compendium of Chromatin Contact Maps Reveal Spatially Active Regions in the Human Genome. Cell Rep 17, 2042 (2016).

88. Mosquera, J. V. et al. Integrative single-cell meta-analysis reveals disease-relevant vascular cell states and markers in human atherosclerosis. (2023) doi:10.1016/j.celrep.2023.113380.

89. Granja, J. M. et al. ArchR is a scalable software package for integrative single-cell chromatin accessibility analysis. Nature Genetics 2021 53:3 53, 403–411 (2021).

90. Hafemeister, C. & Satija, R. Normalization and variance stabilization of single-cell RNA-seq data using regularized negative binomial regression. Genome Biol 20, 1–15 (2019).

91. Gaspar, J. M. Improved peak-calling with MACS2. doi:10.1101/496521.

92. Finucane, H. K. et al. Partitioning heritability by functional annotation using genome-wide association summary statistics. Nat Genet 47, 1228–1235 (2015).

93. Hinrichs, A. S. et al. The UCSC Genome Browser Database: update 2006. Nucleic Acids Res 34, (2006).

94. Suzuki, K. et al. Genetic drivers of heterogeneity in type 2 diabetes pathophysiology. Nature 627, 347–357 (2024).

95. Roychowdhury, T. et al. Genome-wide association meta-analysis identifies risk loci for abdominal aortic aneurysm and highlights PCSK9 as a therapeutic target. Nat Genet 55, 1831–1842 (2023).

96. Keaton, J. M. et al. Genome-wide analysis in over 1 million individuals of European ancestry yields improved polygenic risk scores for blood pressure traits. Nat Genet 56, 778–791 (2024).

97. Malik, R. et al. Multiancestry genome-wide association study of 520,000 subjects identifies 32 loci associated with stroke and stroke subtypes. Nat Genet 50, 524–537 (2018).

98. Murphy, A. E., Schilder, B. M. & Skene, N. G. MungeSumstats: a Bioconductor package for the standardization and quality control of many GWAS summary statistics. Bioinformatics 37, 4593–4596 (2021).

99. Das, S. et al. Next-generation genotype imputation service and methods. Nat Genet 48, 1284–1287 (2016).

100. Loh, P. R. et al. Reference-based phasing using the Haplotype Reference Consortium panel. Nat Genet 48, 1443–1448 (2016).

101. Maples, B. K., Gravel, S., Kenny, E. E. & Bustamante, C. D. RFMix: a discriminative modeling approach for rapid and robust local-ancestry inference. Am J Hum Genet 93, 278–288 (2013).

102. Auton, A. et al. A global reference for human genetic variation. Nature 526, 68–74 (2015).

103. Benaglio, P. et al. Mapping genetic effects on cell type-specific chromatin accessibility and annotating complex immune trait variants using single nucleus ATAC-seq in peripheral blood. PLoS Genet 19, (2023).

104. Munz, M. et al. Qtlizer: comprehensive QTL annotation of GWAS results. Sci Rep 10, (2020).

105. Zhou, W. et al. Efficient and accurate mixed model association tool for single-cell eQTL analysis. medRxiv (2024) doi:10.1101/2024.05.15.24307317.

106. Fulco, C. P. et al. Activity-by-contact model of enhancer–promoter regulation from thousands of CRISPR perturbations. Nature Genetics 2019 51:12 51, 1664–1669 (2019).

107. Ewels, P. A. et al. The nf-core framework for community-curated bioinformatics pipelines. Nat Biotechnol 38, 276–278 (2020).

108. Servant, N. et al. HiC-Pro: an optimized and flexible pipeline for Hi-C data processing. Genome Biol 16, (2015).

109. Marchal, C., Singh, N., Corso-Díaz, X. & Swaroop, A. HiCRes: a computational method to estimate and predict the genomic resolution of Hi-C libraries. Nucleic Acids Res 50, E35 (2022).

110. Abdennur, N. et al. Cooltools: Enabling high-resolution Hi-C analysis in Python. PLoS Comput Biol 20, (2024).

111. Rao, S. S. P. et al. A 3D map of the human genome at kilobase resolution reveals principles of chromatin looping. Cell 159, 1665–1680 (2014).

112. Kaul, A., Bhattacharyya, S. & Ay, F. Identifying statistically significant chromatin contacts from Hi-C data with FitHiC2. Nat Protoc 15, 991–1012 (2020).

113. Xu, W. et al. CoolBox: a flexible toolkit for visual analysis of genomics data. BMC Bioinformatics 22, (2021).

114. Clauset, A., Newman, M. E. J. & Moore, C. Finding community structure in very large networks. (2004).

115. Bioconductor - RBGL. https://bioconductor.org/packages/release/bioc/html/RBGL.html.

116. Anene-Nzelu, C. G. et al. Assigning Distal Genomic Enhancers to Cardiac Disease-Causing Genes. Circulation 142, 910–912 (2020).

117. Wingett, S. et al. HiCUP: Pipeline for mapping and processing Hi-C data. F1000Res 4, (2015).

118. Durand, N. C. et al. Juicer Provides a One-Click System for Analyzing Loop-Resolution Hi-C Experiments. Cell Syst 3, 95–98 (2016).

119. Bhattacharyya, S., Chandra, V., Vijayanand, P. & Ay, F. Identification of significant chromatin contacts from HiChIP data by FitHiChIP. Nat Commun 10, (2019).

120. Tan, W. L. W. et al. Epigenomes of Human Hearts Reveal New Genetic Variants Relevant for Cardiac Disease and Phenotype. Circ Res 127, 761–777 (2020).

121. Li, H. & Durbin, R. Fast and accurate short read alignment with Burrows-Wheeler transform. Bioinformatics 25, 1754–1760 (2009).

122. Kumar, V. et al. Uniform, optimal signal processing of mapped deep-sequencing data. Nat Biotechnol 31, 615–622 (2013).

123. Xie, S., Duan, J., Li, B., Zhou, P. & Hon, G. C. Multiplexed Engineering and Analysis of Combinatorial Enhancer Activity in Single Cells. Mol Cell 66, 285–299.e5 (2017).

124. Sanjana, N. E., Shalem, O. & Zhang, F. Improved vectors and genome-wide libraries for CRISPR screening. Nat Methods 11, 783–784 (2014).

